# Application of a new highly multiplexed amplicon sequencing tool to evaluate *Plasmodium falciparum* antimalarial resistance and relatedness in individual and pooled samples from Dschang, Cameroon

**DOI:** 10.1101/2024.10.03.24314715

**Authors:** Jacob M. Sadler, Alfred Simkin, Valery P. K. Tchuenkam, Isabela Gerdes Gyuricza, Abebe A. Fola, Kevin Wamae, Ashenafi Assefa, Karamoko Niaré, Kyaw Thwai, Samuel J. White, William J. Moss, Rhoel R. Dinglasan, Sandrine Nsango, Christopher B. Tume, Jonathan B. Parr, Innocent Mbulli Ali, Jeffrey A. Bailey, Jonathan J. Juliano

## Abstract

**Background:** Resistance to antimalarial drugs remains a major obstacle to malaria elimination. Multiplexed, targeted amplicon sequencing is being adopted for surveilling resistance and dissecting the genetics of complex malaria infections. Moreover, genotyping of parasites and detection of molecular markers drug resistance in resource-limited regions requires open-source protocols for processing samples, using accessible reagents, and rapid methods for processing numerous samples including pooled sequencing.

**Methods:** *<u>P</u>lasmodium falciparum* <u>S</u>treamlined <u>M</u>ultiplex <u>A</u>ntimalarial <u>R</u>esistance and <u>R</u>elatedness <u>T</u>esting (*Pf*-SMARRT) is a PCR-based amplicon panel consisting of 15 amplicons targeting antimalarial resistance mutations and 9 amplicons targeting hypervariable regions. This assay uses oligonucleotide primers in two pools and a non-proprietary library and barcoding approach.

**Results:** We evaluated *Pf*-SMARRT using control mocked dried blood spots (DBS) at varying levels of parasitemia and a mixture of 3D7 and Dd2 strains at known frequencies, showing the ability to genotype at low parasite density and recall within-sample allele frequencies. We then piloted *Pf*-SMARRT to genotype 100 parasite isolates collected from uncomplicated malaria cases at three health facilities in Dschang, Western Cameroon. Antimalarial resistance genotyping showed high levels of sulfadoxine-pyrimethamine resistance mutations, including 31% prevalence of the DHPS A613S mutation. No K13 candidate or validated artemisinin partial resistance mutations were detected, but one low-level non-synonymous change was observed. *Pf*-SMARRT’s hypervariable targets, used to assess complexity of infections and parasite diversity and relatedness, showed similar levels and patterns compared to molecular inversion probe (MIP) sequencing. While there was strong concordance of antimalarial resistance mutations between individual samples and pools, low-frequency variants in the pooled samples were often missed.

**Conclusion:** Overall, *Pf*-SMARRT is a robust tool for assessing parasite relatedness and antimalarial drug resistance markers from both individual and pooled samples. Control samples support that accurate genotyping as low as 1 parasite per microliter is routinely possible.

**SCOPE STATEMENT (200):** Malaria remains a critical global public health problem. Antimalarial drug resistance has repeatedly undermined control and the emergence of artemisinin partial resistance in Africa is the latest major challenge. Malaria molecular surveillance (MMS) has emerged as a powerful tool to monitor molecular markers of resistance and changes in the parasite population. Streamlined methods are needed that can be readily adopted in endemic countries. We developed *<u>P</u>lasmodium falciparum* <u>S</u>treamlined <u>M</u>ultiplex <u>A</u>ntimalarial <u>R</u>esistance and <u>R</u>elatedness <u>T</u>esting (*Pf*-SMARRT), a multiplex amplicon deep sequencing approach that uses easily accessible products without proprietary steps and can be sequenced on any Illumina sequencer. We validated this tool using controls, including mocked dried blood spots, and then implemented it to evaluate resistance and parasite relatedness among 100 samples from Cameroon. The assay was able to reliably assess the within-sample allele frequency of antimalarial resistance markers and discriminate strains within and between individuals. We also evaluated a more cost-effective surveillance approach for antimalarial resistance polymorphisms using pooled samples. While within-pool frequencies of mutations were accurate in pools with higher numbers of samples, this resulted in the loss of the ability to detect variants uncommon in the pool. Overall *Pf-*SMARRT provides a new protocol for conducting MMS that is easily implementable in Africa.

## INTRODUCTION

Increasing resistance to antimalarial drugs remains a major hurdle for malaria control programs. Despite some gains over the last decades, malaria control has flattened if not reversed in some parts of Africa. Globally, malaria resulted in 249 million infections and 608,000 deaths in 2022 (World Health Organization, 2022). The recent emergence of partial artemisinin resistance in Africa (Rosenthal et al., 2024) is of particular concern considering the clinical failure of artemisinin combination therapies (ACTs) observed in Southeast Asia (Dondorp et al., 2009; Noedl et al., 2010). Antimalarial efficacy has traditionally been monitored through *in vivo* efficacy studies, often termed therapeutic efficacy studies (TES), and by *in vitro* measurement of response to antimalarials (Ekland and Fidock, 2008; White, 2022). However, once correlated mutations are discovered across large populations, antimalarial resistance can be more efficiently tracked by measuring individual mutations with malaria molecular surveillance (MMS) (Prosser et al., 2018; Juliano et al., 2024). Given that critical mutations occur in multiple locations in multiple genes, simple and rapid methods for assessing numerous known mutations are needed. One approach that has become more popular in recent years is targeted deep sequencing through PCR amplicons or molecular inversion probes (MIPs) (Aydemir et al., 2018; Kattenberg et al., 2023; Aranda-Díaz et al., 2024). Multiplex targeted deep sequencing can also help to understand how parasites are related to one another. By targeting single nucleotide polymorphisms (SNPs) or microhaplotypes, multiplex assays can be used to study overall population diversity, within-individual diversity, or even to track transmission by leveraging metrics like identity-by-descent (IBD).

Multiplex targeted sequencing can represent direct amplification (multiplex PCR) or initial capture enrichment followed by universal PCR. MIPs allow for high levels of multiplexing (100s-1000s of targets) and are rapidly adaptable. However, MIPs are not as sensitive at low levels of parasitemia compared to smaller multiplex PCR panels (often 10 to 100 targets).

Amplicon panels, in contrast, can be challenging to multiplex and balance successfully, making them less adaptable. They are often lower throughput, with multiple PCR reactions required to limit cross amplification between products. Many PCR amplicon panels have been developed that target antimalarial resistance markers, parasite diversity, or both (LaVerriere et al., 2022; Holzschuh et al., 2023; Rovira-Vallbona et al., 2023; Aranda-Díaz et al., 2024). These panels have mostly focused on short amplicons but recently, longer amplicons have become accessible using Oxford Nanopore Technology (Girgis et al., 2023; de Cesare et al., 2024). For an amplicon panel to be widely useful, an assay must be highly sensitive in genotyping low-parasitemia samples, capable of high-throughput use, low cost, and easily implemented in resource-limited areas. One way to achieve these goals is to design approaches for which materials are easily acquired through multiple commercial sources and detailed protocols are publicly available.

When surveilling for mutations, one way to make MMS more efficient is the use of pooled sequencing, for which individual samples or DNA extracts are mixed prior to amplification. Pooled-sample sequencing was piloted in malaria parasites but usually by using a single amplicon at a time (Taylor et al., 2013, 2015; Juliano et al., 2016; Brazeau et al., 2019). This approach shows promise, providing frequency estimates in the pools similar to prevalence estimates of the individual samples (Taylor et al., 2013). However, the reliability of reported estimates and the application of pooling to highly multiplexed amplicon sequencing is yet to be established. Success of pooled sequencing using molecular inversion probes (MIPs) suggests that multiplex amplicon sequencing should be feasible using pooled samples (Aydemir et al., 2021; Young et al., 2024).

Studies assaying molecular markers of antimalarial resistance have become more common in Cameroon (**Table 1**). Overall, mutations associated with sulfadoxine-pyrimethamine resistance in *Plasmodium falciparum* dihydrofolate reductase (DHFR) and dihydropteroate synthase (DHPS) are often reported (Chauvin et al., 2015). The K76T mutation in *P. falciparum* chloroquine resistance transporter (CRT) is no longer commonly seen (Niba et al., 2023).

**Table 1.**
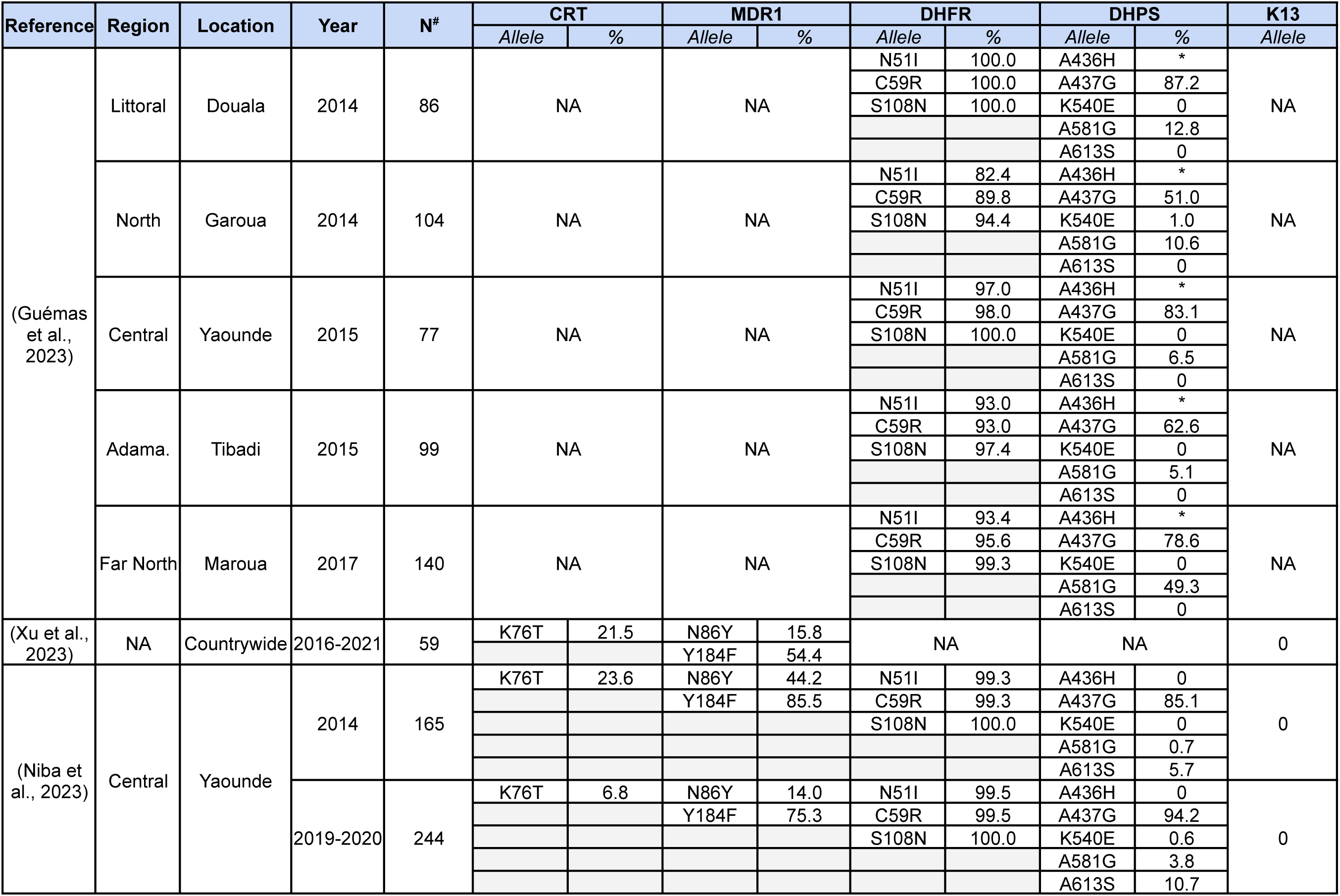
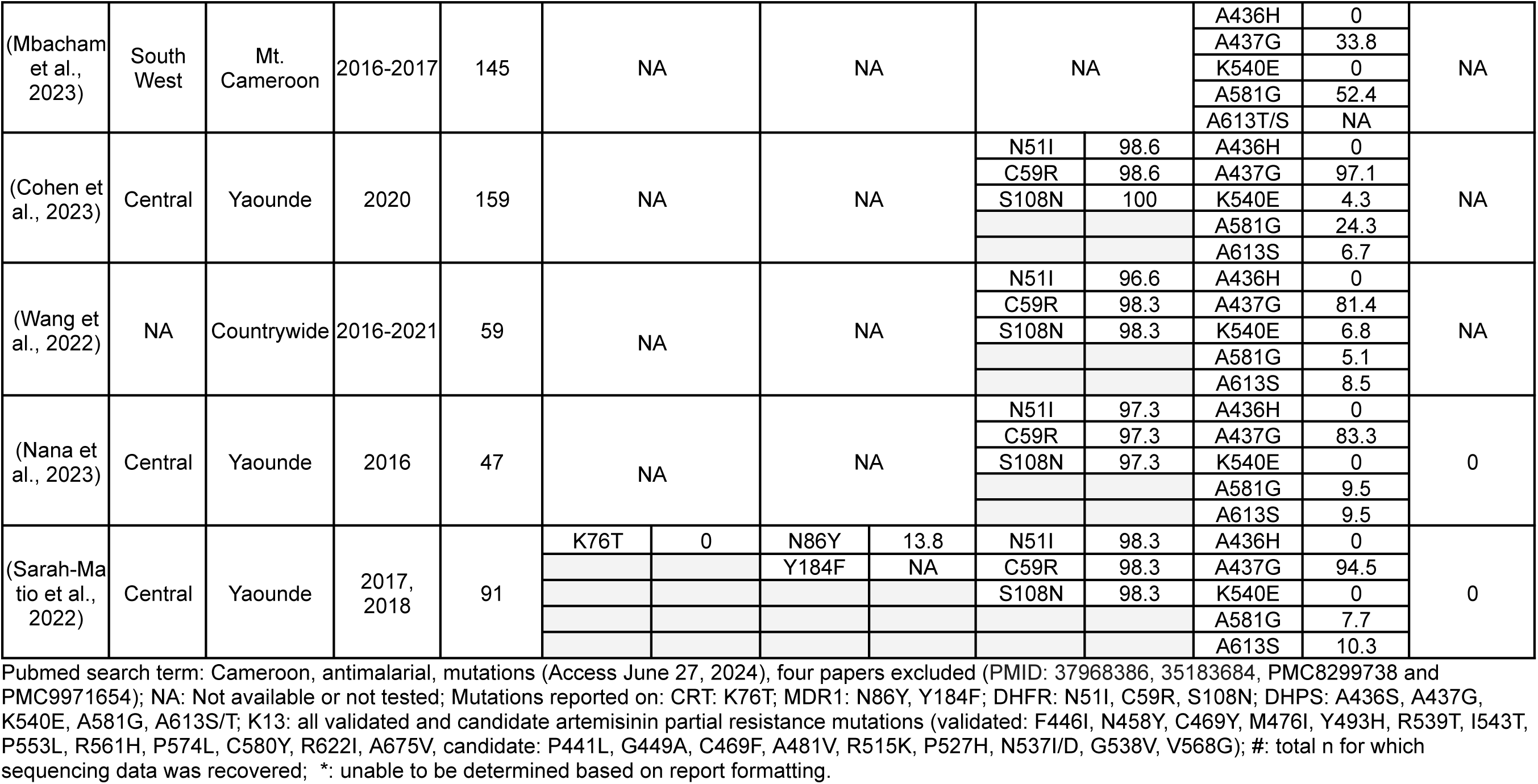
Recent reports (published 220-2024) of molecular markers of *P. falciparum* antimalarial resistance mutations in Cameroon.

Mutations in *P. falciparum* multidrug resistance protein 1 (MDR1) are less common, with the N86Y mutation frequently identified in studies that sequenced the *mdr1* gene (Tuedom et al., 2021). To date, validated and candidate artemisinin partial resistance polymorphisms in *P. falciparum* kelch13 (K13) have not been described in Cameroon (Niba et al., 2023).

Here, we present a new highly multiplexed amplicon panel for studying antimalarial resistance and parasite relatedness, ***<u>P</u>****lasmodium **f**alciparum* **<u>S</u>**treamlined **<u>M</u>**ultiplex **<u>A</u>**ntimalarial **<u>R</u>**esistance and **<u>R</u>**elatedness **<u>T</u>**esting (***Pf*-SMARRT**). This protocol uses commercially available reagents that can be sourced from multiple vendors using any multiplex capable Taq polymerase. We assessed assay characteristics using mocked blood spots at varying parasitemias and a known mixture of two malaria strains. The assay was then used to characterize antimalarial resistance polymorphisms, complexity of infection (COI), and genetic relatedness of 100 *P. falciparum* infections from Dschang, Cameroon. We compared the assay performance against MIPs on a subset of 50 samples to examine how the assay performs against a routinely implemented method in our laboratories (Verity et al., 2020). We also evaluated the performance of the assay in detecting and quantifying parasite genetic variation using pooled samples.

## METHODS

### Development of the *<u>P</u>lasmodium falciparum* <u>S</u>treamlined <u>M</u>ultiplex <u>A</u>ntimalarial <u>R</u>esistance and <u>R</u>elatedness <u>T</u>esting (*Pf*-SMARRT) assay

By leveraging previously published and validated primer pairs (**Supplemental Protocol 1**), as well as development of specific primers for the assay, we targeted 15 antimalarial resistance polymorphisms, as well as 9 regions of hypervariability that don’t contain any length polymorphisms. This resulted in a panel containing 24 target amplicons. The major antimalarial resistance polymorphisms targeted by the assay include:

● K13: amino acids 433-570, 575-623 and 668-702; covering almost all current validated (F446I, N458Y, C469Y, M476I, Y493H, R539T, I543T, P553L, R561H, C580Y, R622I, A675V) and candidate (P441L, G449A, C469F, A481V, R515K, P527H, N537I/D, G538V, V568G) artemisinin partial resistance mutations.
● CRT: amino acids 43-90; including K76T
● MDR1: amino acids 71-92, 170-229, and 1031-1042; including N86Y, Y184F, and S1034C
● DHFR: amino acids 41-81 and 97-123; including, N51I, C59R, and S108N
● DHPS: amino acids 429-443, 537-573, and 579-615; including I431V, A436S, A437G, K540E, A581G, and A613S/T

Multiplex amplification was performed in two reactions (Primer Pool A and B) to avoid overlapping amplicons. Post-amplification, the PCR products were purified using silica micro columns. Pool A and B were quantified using Qubit HS dsDNA reagent, mixed at equal concentrations, then prepared into libraries for sequencing using a ligation-based protocol (Glenn et al., 2019). The complete protocol for sample preparation is provided in **Supplemental Protocols 1** and **2**. After sequencing using Illumina 2 X 150bp chemistry, sample haplotypes and their within-sample frequency were determined using `SeekDeep` (Hathaway et al., 2018). Samples were sequenced with at least 2 replicates allowing for improved de-noising. `SeekDeep` can examine each replicate individually, limiting erroneous haplotypes with depth and frequency filters. Replicates can be combined into a final sample call, which offers the same depth and frequency filters but requires haplotypes to be in both replicates, thereby minimizing false haplotypes and more accurately estimating within-sample haplotype frequency.

### Assay performance and validation of SeekDeep processing filters using control data

To validate the assay, DNA from the mocked blood spots at varying levels of parasitemia (parasites per microliter: p/µL) (10,000 p/µL, 1,000 p/µL, 100 p/µL, 10 p/µL, and 1 p/µL) was amplified and sequenced twelve times using two replicates per sample. In addition, a mixture containing 88% 3D7 and 12% Dd2 genomic DNA (MRA-102G and MRA-150G, respectively; BEI Resources, Manassas, VA) at a concentration of 1,000 p/µL was sequenced twelve times with two replicates per sample. Final `SeekDeep` sample filtering was: 1) a minimum of 100 reads per sample; 2) a minimum of 100 reads per replicate; 3) amplicons were restricted to the exact expected length; 4) a minimum frequency within a sample for a haplotype of 1%; and 5) a requirement that haplotypes needed to pass thresholds in both replicates to be counted.

### Clinical samples and ethics

1. *P. falciparum* positive samples from an acute febrile illness cohort conducted in Dschang, Cameroon from June 12, 2020 –September 8, 2020 were used for this study. The cohort has been described previously (Ali et al., 2022). Briefly, 431 participants presenting with fever (axillary temperature ≥37.5°C or self-reported history of fever) in the past 24 hours without signs and symptoms of severe malaria to Dschang Regional Hospital, Hospital de Soeur Servantes du Christ de Batsingla, and Saint Vincent Hospital outpatient departments. Individuals were excluded if they had used anti-malarials in the past 14 days. Consented participants provided dried blood spots (DBS) collected via finger prick. Demographic data was collected. The study was approved by the institutional review board (IRB) of the Cameroon Baptist Convention Health Board (FWA00002077), Protocol IRB2019-40.(Ali et al., 2022) Written informed consent was administered in French or English, based on participant preference, via an independent translator (who also spoke the local language Yemba).(Ali et al., 2022) For children, informed consent was obtained from a parent or guardian and assent from adolescents. Molecular analysis of de-identified samples and data were deemed non-human subjects research by the University of North Carolina IRB.

### Real-time PCR for estimating parasitemia

One hundred samples with an estimated parasite density of 100 parasites/µL or greater were randomly selected from a set of previously generated data using primers targeting *varats,* a *P. falciparum* specific multi-copy target (Hofmann et al., 2015) (**Figure S1**). All 100 samples were re-extracted from 3 X 6 mm DBS punches using a Chelex resin extraction (Popkin-Hall et al., 2024). Parasite density in re-extracted DNA was confirmed by real-time PCR targeting *varats,* as previously described (Hofmann et al., 2015). To create a standard curve and determine estimated parasitemia for each sample, standard dilutions of Chelex-extracted, mocked DBS containing whole blood spiked with known densities of cultured 3D7 *P. falciparum* genomic DNA (MRA-102, BEI Resources, Manasas, VA) were amplified alongside clinical samples. The resulting Ct values of the standard dilutions were plotted against known parasite densities to form a standard curve, then each sample’s Ct was plotted on the standard curve to calculate each sample’s parasitemia. All runs included non-template control reactions. Samples with low estimated parasitemia on initial real-time PCR runs, when re-extracted, were checked for DNA concentration using a Qubit fluorometer (Invitrogen, Carlsbad, California) and Qubit 1X HS dsDNA quantification reagent to ensure proper extraction occurred.

### Individual clinical sample sequencing

The workflow of clinical samples is shown in **Figure 1**. The 100 samples were sequenced with two replicates. To evaluate initial amplification and determine if additional sequencing was needed, we assessed each replicate separately using less stringent filtering criteria. These filters were set to be as permissive as possible and required only a minimum of 25 reads per replicate. Using these criteria, samples were either: 1) considered complete (if the same haplotypes were found in both replicates and the difference in haplotype frequency between replicates was never greater than 10%); or 2) were re-amplified and resequenced with two new replicates (different haplotypes detected between replicates and/or >10% variation in haplotype frequency found between replicates for any haplotype). The replicate PCRs of one sample still showed an increase in the number of haplotypes with >10% variation in haplotype frequency upon resequencing, and the original replicates for that sample were retained. After repeat sequencing for samples requiring new replicates, the final data was called at the sample level using the same filters applied to the controls.

**Figure 1.**
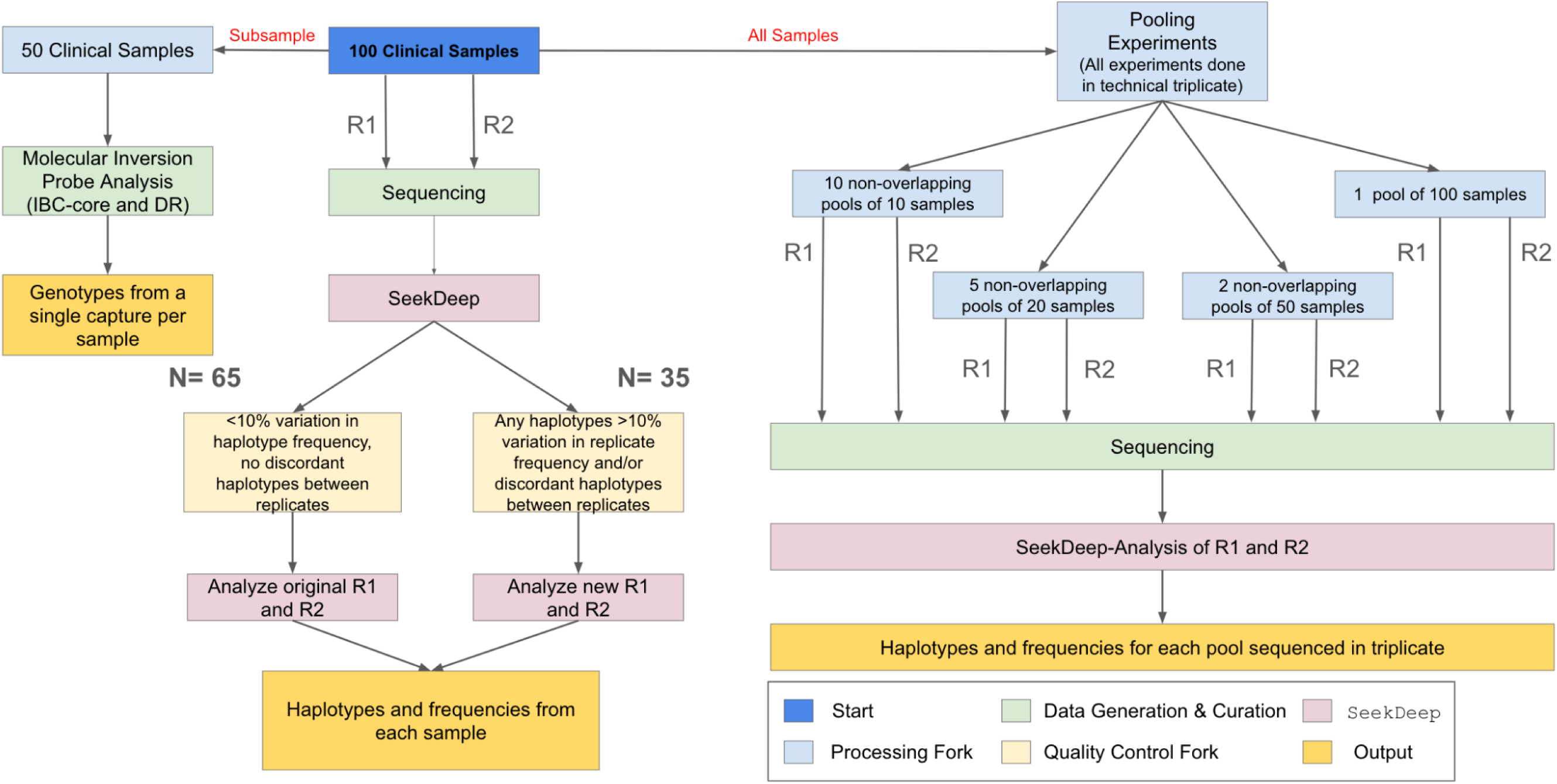
**Sample flow**. One hundred samples from parasitemic participants were used in the study. These samples were PCRed and sequenced with replicates (R1 and R2) and processed in `SeekDeep`. To ensure high quality field sample data, we examined the replicates for each field sample. Replicates that differed in the haplotypes reported or that showed >10% variation in haplotype frequency between replicates (N=35) were re-amplified and re-sequenced, with one exception. In addition, a subset of 50 samples also underwent MIP sequencing to allow for comparison of relatedness and antimalarial resistance frequency. Lastly, the 100 samples were used to generate non-overlapping pools of N=10, N=20, N=50, and N=100 to evaluate the utility of the assays for pooled sequencing. All final sample (and pool) genotyping calls are based upon haplotypes that occurred in both replicates (R1 and R2) and within sample haplotype frequency was averaged between replicates.

### Molecular inversion probe (MIP) sequencing

A subset of 50 isolates were selected for MIP sequencing. Samples were captured and sequenced as previously described using capture probes for genome-wide single nucleotide polymorphisms and probes targeting known antimalarial resistance polymorphisms (Verity et al., 2020). Data was processed using MIP analyzer as previously described (Verity et al., 2020). A variant required at least 3 unique molecular identifiers to be called and a sample required a depth of 10 UMIs.

### Pooled sample sequencing

Pools of samples were generated with a range of pool sizes. The number of pools available at each sample size was a function of the available number of nonoverlapping groups of samples. Thus we generated ten pools of 10 samples, five pools of 20 samples, two pools of 50 samples, and one pool of 100 samples. Each pool was sequenced in triplicate with two replicates informing each pool. The pools were analyzed in SeekDeep with the same clustering parameters as the individual samples, except that in the pooled samples the minimum frequency within a sample for a haplotype was set to 0.5%, on the assumption that more samples should give more power to confidently call low frequency haplotypes.

### Data Analysis

For comparisons of study participants, p-value comparisons across groups for categorical variables are based on the chi-square test of homogeneity, whereas p-values for continuous variables are based on the Kruskal-Wallis test for median.

Drug resistance and other genomic metrics for both *Pf*-SMARRT and MIP data were calculated using the same approach. Drug resistance prevalence was determined by considering both heterozygous and homozygous mutant alleles as mutant samples, while homozygous reference alleles were considered wild-type samples. Loci that were not sequenced within a sample were not included for prevalence calculation (p=x/n*100), where p = prevalence, x = number of mutant alleles, n = number of successfully genotyped loci (Fola et al., 2023). The n varied across loci due to differences in sequence coverage for each amplicon. The complexity of infection (COI) was estimated in two ways: 1) a counting method where the amplicon with the highest number of haplotypes was considered the sample COI and 2) using SNP data from the amplicons to assess COI using `THE REAL McCOIL` (Chang et al., 2017). The program was run in the categorical method format: heterozygous call as 0.5, homozygous reference allele as 0, homozygous alternative allele as 1 and no call as −1. Heterozygosity (He) for each amplicon, the probability that two amplicon haplotypes will be different, was determined as 1-(p_1_^2^+p_2_^2^ +p_n_^2^), where p_1_, p_2_, etc. are the population frequencies of each haplotype and n is the total number of haplotypes in the population.The ability to discern unrelated strains, in that any of the amplicons would have different haplotypes, was estimated as 1-((1-He_1_)*(1-He_2_)* (1-He_i_)) where He_1_, He_2_, etc. are the heterozygosities at each amplicon and i is the total number of amplicons. To assess whether parasite populations within Cameroon clustered by their geographic origin, we conducted principal component analysis (PCA) using `SNPRelate` function in R 4.2.1 software. The results were visualized using `ggplot2` or GraphPad Prism (v.10.2.3).

## RESULTS

### Assay performance at different parasitemias in monoclonal controls

Sequencing depth for test validation controls are shown in **Figure S2**. In general, replicate samples had very similar read counts at all parasitemias except 1 p/µL, where more variation was introduced in part due to loci failing more commonly at that level of parasitemia. Genotyping was highly successful at most of the amplicons for parasitemias as low as 10 p/µL, with some amplicons dropping out at 1 p/µL (mean 19.16 amplicons, range: 9-24) (**Figure S3**). The results showed the estimated number of haplotypes expected, with a single haplotype reported in most conditions across parasitemia levels and some samples with no called haplotypes at the lowest parasitemias (**Figure S4** for diversity markers and **Figure S5** for antimalarial resistance markers).

Unexpected (false-positive haplotypes that should not be identified in 3D7) occurred in the DHPS-540 amplicon in two samples at 1 p/µL and in the Heome-A amplicon in two samples at 1 p/µL. This represented 4 of 1,440 amplicons (24 amplicons across 12 samples across 5 parasitemias) that had false positive calls, and these occurred only at the lowest parasitemia.

These unexpected haplotypes were low in frequency (<6%) (**Figure S6**). The issue of low frequency false-positive haplotypes has been described previously with other amplicon panels (Mideo et al., 2016; Early et al., 2019).

### Estimation of within-sample haplotype frequency in the control mixture

Upon testing the 3D7/Dd2 mixture (ratio 88:12), we observed high concordance between measured and expected allele frequency for the diversity markers and antimalarial SNPs (**Figure S7** and **Table S2**) with one exception. The DHPS 436/437 amplicon consistently underestimated the Dd2 SNP frequency when detected (mean 1.09%, SD 0.11%), and often failed to detect it (7/12). These controls were run separately from the clinical samples, which routinely detected multiple haplotypes in the clinical samples. Thus, it is unclear why this amplicon failed in the control reactions. MDR1 SNP frequencies varied from expected due to the copy number variation in Dd2, which contains different point mutations in each copy.

### Cameroon participant characteristics

A total of 230 of the 431 participants were positive for *P. falciparum* by real-time PCR. Clinical characteristics of malaria positive study participants are summarized in **Table S1**. The participants were 54% female and their average age was 26 years (range: 20-40) and most resided in Dschang (79%). The majority had used a bed net the previous night (57%). Prior treatment was not an exclusion and most had not been treated for malaria in the last 14 days (74%). The distribution of estimated parasitemias is shown in **Figure S1**. The 100 participants selected for sequencing did not differ significantly from those that were not selected, or from the total malaria positive population, and had a mean parasitemia of 10,498 p/µL (Range: 133 - 94,560 p/µL). Parasitemias after re-extraction of DBS used for sequencing from the 100 participants had a mean parasitemia of 7103.8 p/µL (Range: 0.5 - 79,627 p/µL). The slightly lower overall parasitemia is likely due to the re-extraction occurring nearly 2 years later than the original from DBS stored at -20C. Once sample appears to have had a poor re-extraction. Individual characteristics of the 100 sequenced participants are shown in **Table S3**.

### Individual sample sequence quality

Initial amplification resulted in 91 of 100 samples in which the haplotype sequences were the same in both replicates. Sixty-five of these samples had the same haplotype sequences, and haplotype frequencies never differed by more than 10% (**Figure 1**). After re-amplifying and resequencing to increase the depth of replicates in the 35 samples that either had haplotypes >10% different in frequency or discordant haplotypes between replicates (**Figure 1**), the final dataset contained 97 samples that passed all filters and 3 samples that had one haplotype with >10% difference in frequencies between replicates. In the final sequence data, most samples were successfully genotyped at nearly all 24 amplicons (mean: 23.7 amplicons genotyped, SD: 0.67, range: 20-24) (**Figure S8**). The within-sample allele frequency between the individual replicates of the same sample showed a high level of concordance (**Figure S9**). Sequencing depth was high, averaging >2^10^ reads/amplicon/sample, across most amplicons, with depth lower for the Heome-H amplicon (**Figure S10**).

### Complexity of infection

The complexity of infection (COI) was first determined simply by the greatest number of haplotypes observed for any amplicon in a sample (**Figure 2A**). The overall mean COI was similar between the 100 sequenced samples (2.7, Range 1-5) and the 50 sample subset that underwent both *Pf-*SMARRT and MIP analysis (2.6, Range 1-5).

**Figure 2:**
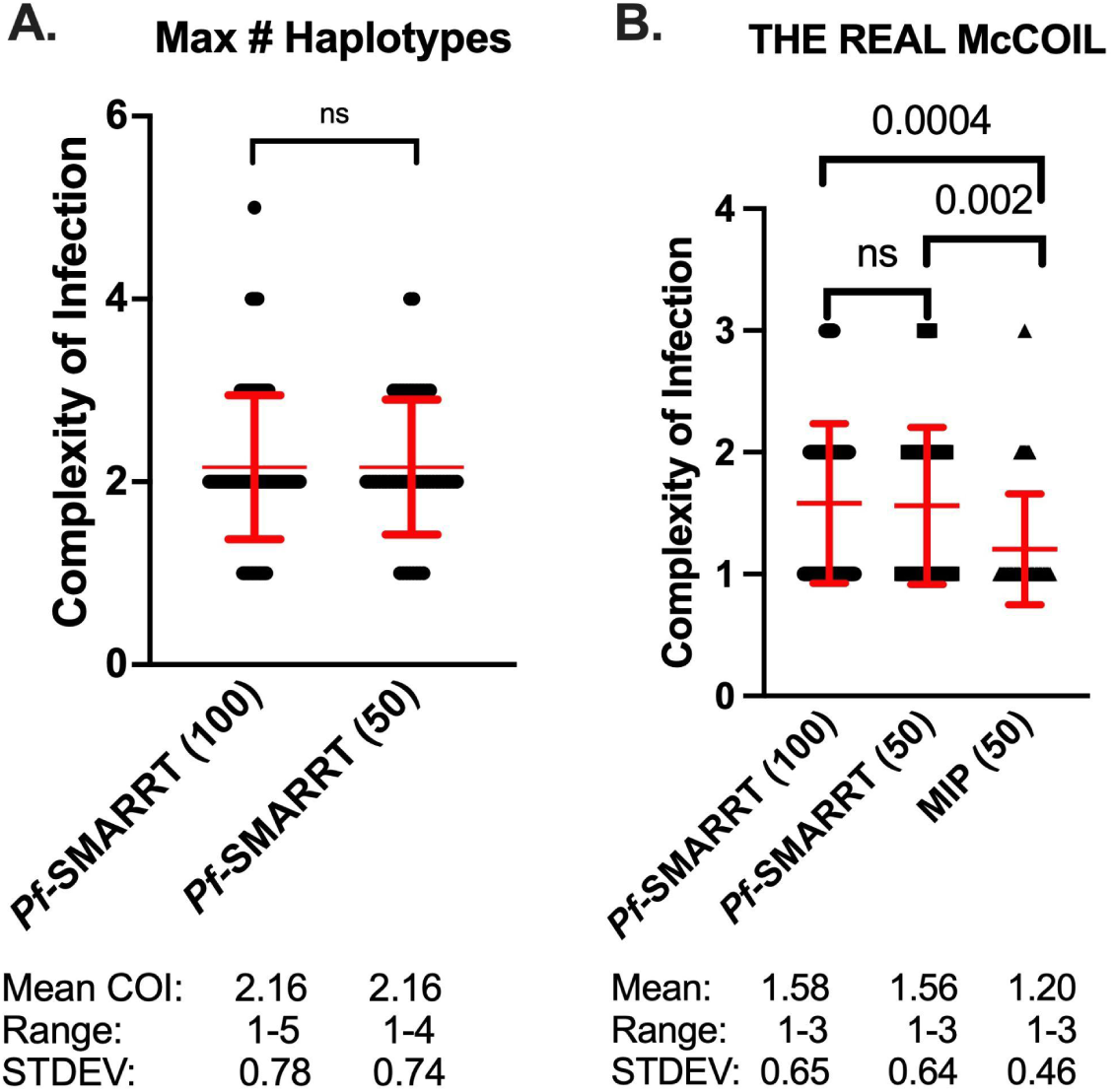
Estimated complexity of infection for each sample. Red text denotes the mean (as a central dot) and standard deviation (as arms above and below the mean). Black dots represent COI values for individual samples. Panel A depicts the COI for *Pf*-SMARRT based on the amplicon that provided the highest number of haplotypes. Panel B shows the estimated COI determined by THE REAL McCOIL for *Pf*-SMARRT and MIPs. Significance determined by t-test.

*Pf*-SMARRT’s absolute count of haplotypes showed a higher mean COI than estimates by `THE REAL McCOIL`. When COI was estimated by `THE REAL McCOIL`, *Pf-*SMARRT estimated significantly higher mean COI than MIP sequencing (figure 2B). The relative COI for each amplicon based on haplotype count of *Pf*-SMARRT are shown in **Figure S11**. There was no single amplicon that was the most reliable indicator of maximum haplotype count. However, Heome-A was the most informative amplicon, being associated with the maximum haplotype count in 50 samples, while Heome-D was associated with the maximum haplotype count in 46 samples, and AMA1 in 43 samples.

### Antimalarial Resistance

Genotyping results and the estimated prevalence of common antimalarial resistance polymorphisms are shown for key resistance genes in **Tables 2** and **3**. No WHO validated or candidate artemisinin partial resistance polymorphisms were found, but one sample with a non-synonymous (A578**S**) and one sample with a synonymous (F522F) mutation were found in K13. Among the 100 samples sequenced, DHFR mutations associated with pyrimethamine resistance were at fixation. DHPS mutations, associated with sulfadoxine resistance, were more variable, while high levels of the DHPS A436**S** and A437**G** mutations, but lower prevalences for the K540**E** (7.0, 2.0-12.3), A581**G** (24.0, 15.6-32.4) and A613**S** (31.0, 21.6-40.1). The majority of infections were wild type at MDR1 amino acid **N**86Y (94.0, 92.13-96.8), a mutation associated with decreased sensitivity to lumefantrine. When compared to MIP sequencing, *Pf*-SMARRT generally identified a higher prevalence of antimalarial resistance polymorphisms (**Table 3**). In part, this appears to be associated with a higher rate of finding mixed genotype infections. *Pf*-SMARRT identified 12.5% (81/650) loci successfully genotyped as mixed genotypes, whereas MIPs found only 6.4% (40/628) (**Table 2**). Detailed data on coverage and allele calls is available in **Table S3**.

**Table 2.** Antimalarial resistance infection genotypes comparing *Pf*-SMARRT and MIPs.

| Gene | Mutation | Full sample set (N=100) |  |  |  | Sub-sample (N=50) |  |  |  |  |  |  |  |
| --- | --- | --- | --- | --- | --- | --- | --- | --- | --- | --- | --- | --- | --- |
|  |  | *Pf*-SMARRT |  |  |  | *Pf*-SMARRT |  |  |  | MIP |  |  |  |
|  |  | n | WT | Mixed | MT | n | WT | Mixed | MT | n | WT | Mixed | MT |
| CRT | K76T | 100 | 81 | 11 | 8 | 50 | 37 | 6 | 7 | 48 | 43 | 1 | 4 |
| MDR1 | N86Y | 100 | 94 | 5 | 1 | 50 | 47 | 2 | 1 | 49 | 47 | 0 | 2 |
|  | Y184F | 100 | 28 | 25 | 47 | 50 | 14 | 13 | 23 | 47 | 20 | 2 | 25 |
| DHFR | N51I | 100 | 0 | 0 | 100 | 50 | 0 | 0 | 50 | 48 | 0 | 1 | 47 |
|  | C59R | 100 | 0 | 0 | 100 | 50 | 0 | 0 | 50 | 48 | 0 | 1 | 47 |
|  | S108N | 100 | 0 | 0 | 100 | 50 | 0 | 0 | 50 | 49 | 0 | 0 | 49 |
| DHPS | I431V | 100 | 62 | 21 | 17 | 50 | 27 | 13 | 10 | 48 | 34 | 3 | 11 |
|  | A436S | 100 | 27 | 32 | 41 | 50 | 14 | 17 | 19 | 48 | 24 | 24 | 0 |
|  | A437G | 100 | 3 | 7 | 90 | 50 | 2 | 1 | 47 | 48 | 5 | 1 | 42 |
|  | K540E | 99 | 92 | 7 | 0 | 49 | 46 | 3 | 0 | 48 | 46 | 1 | 1 |
|  | A581G | 100 | 76 | 21 | 3 | 50 | 37 | 12 | 1 | 49 | 35 | 3 | 11 |
|  | A613S | 100 | 69 | 24 | 7 | 50 | 33 | 14 | 3 | 49 | 34 | 3 | 12 |
| K13 | Multiple* | 100 | 100 | 0 | 0 | 50 | 50 | 0 | 0 | 49 | 49 | 0 | 0 |

n: number of successfully sequenced samples; WT: pure wild type sensitive genotype; Mixed: mixed wild type and resistant genotype; MT: pure mutant genotype; Prev: prevalence of infections containing resistant genotypes; \*: reporting only WHO validated and candidate K13 mutations. MIP: molecular inversion probe.

**Table 3.**
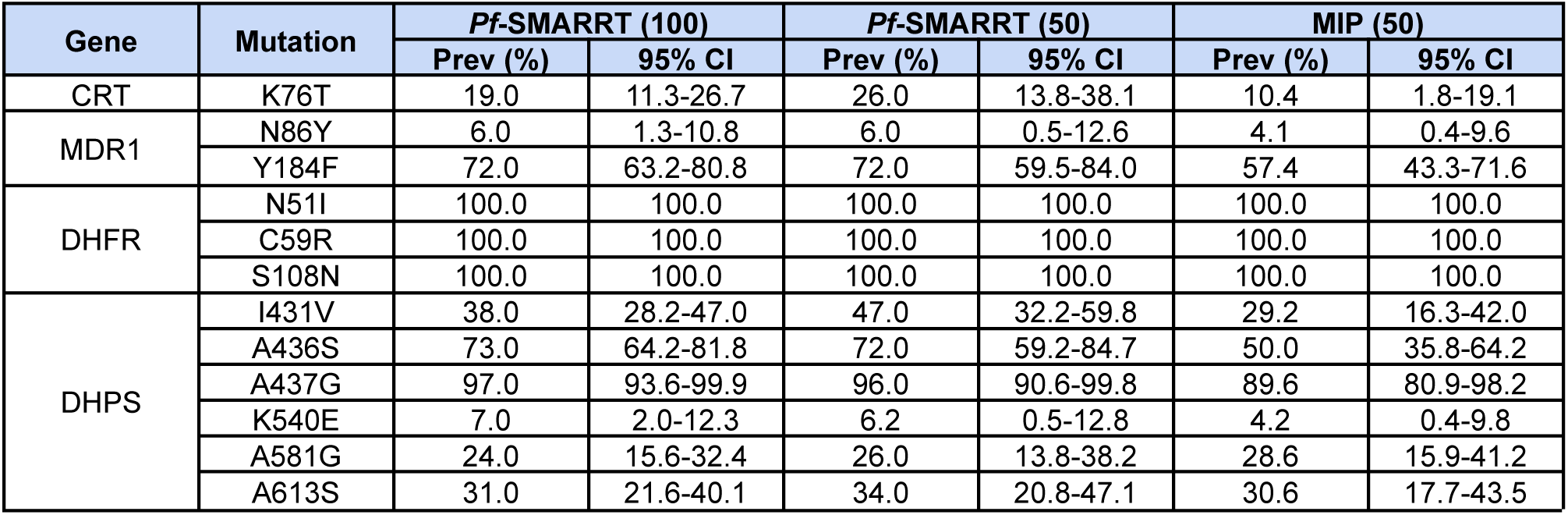
Prevalence of antimalarial resistance polymorphisms by *Pf*-SMARRT vs MIP.

### Parasite diversity

We investigated parasite population structure using *Pf*-SMARRT. Given the majority of samples were drawn from essentially one relatively small geographic location (262 sq. km), we would expect tight clustering of the isolates. Overall, *Pf*-SMARRT confirmed this expectation (**Figure 3**), with principal components 1 and 2 (PC1 and PC2) representing only 3.8% and 3.6% of the variation within the population, respectively. The most distant sample, an individual from Douola who came to the local clinic (approximately 215 km by road), segregated from other samples by PC1. To compare the clustering determined by *Pf*-SMARRT relative to a higher density genotyping method, we compared it to 50 samples also genotyped by MIPS (**Figure S12**). As expected, MIPs provided visually more clustered samples than *Pf*-SMARRT.

**Figure 3.**
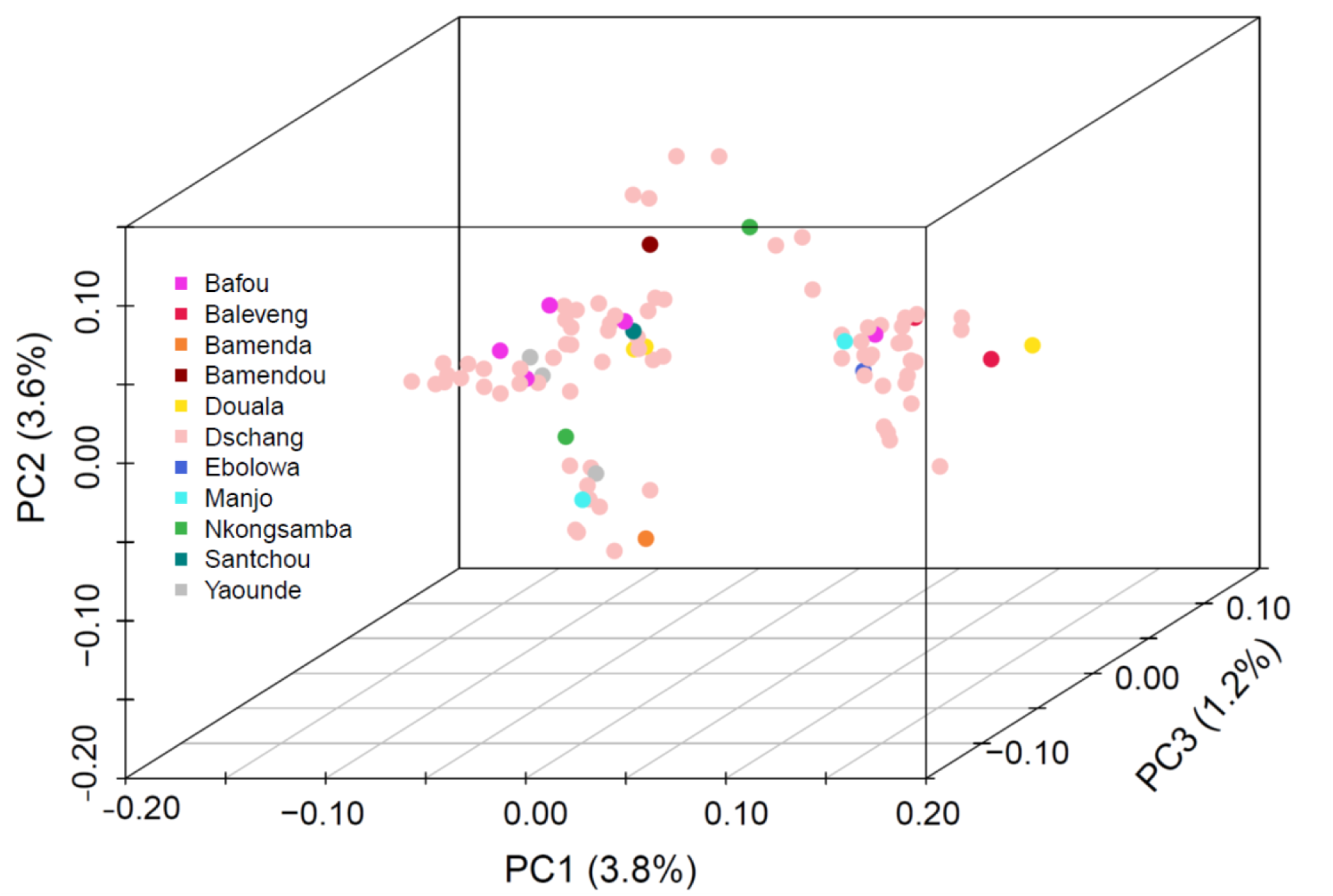
Principal component analysis of 100 Cameroonian samples collected from three hospitals in Dschang, Cameroon. Samples are color coded by the patient’s village/town/city of origin.

Diversity markers will have variable information for discerning strains in different populations. In Cameroon, these diversity markers had a range of heterozygosity from 0.32 to 0.95 (**Table 4**). Unsurprisingly, AMA1 showed the greatest within-population heterozygosity. There were between 5 and 37 haplotypes in the population depending on the loci.

**Table 4.**
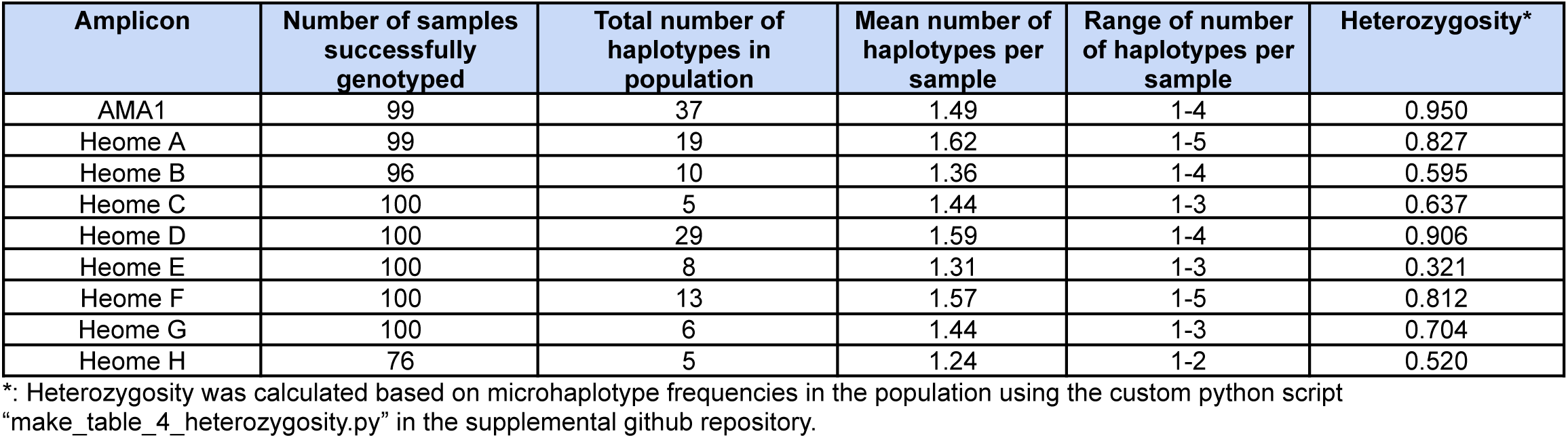
Diversity of amplicon microhaplotypes in the population.

### Estimation of antimalarial resistance frequency using pooled sequencing

Utilizing non-overlapping pools of different sizes, we evaluated the detectability and estimated frequencies of key antimalarial resistance polymorphisms in the pools relative to the weighted (based on parasitemia) frequency based upon individual sample sequencing. In general, correlations between the data were high (**Figure 4**), with more variation in the frequency estimates with smaller pool size. In terms of the ability to detect low abundance alleles, pooling did well for mutations greater than 40% estimated frequency by individual sample sequencing (108/110 detected, 98.2%) (**Figure S13**). However, when a mutation was between 20% and 40% only 22/27 (81.5%) were detected and for polymorphisms less than 20% only 50/79 (63.3%) were detected.

**Figure 4.**
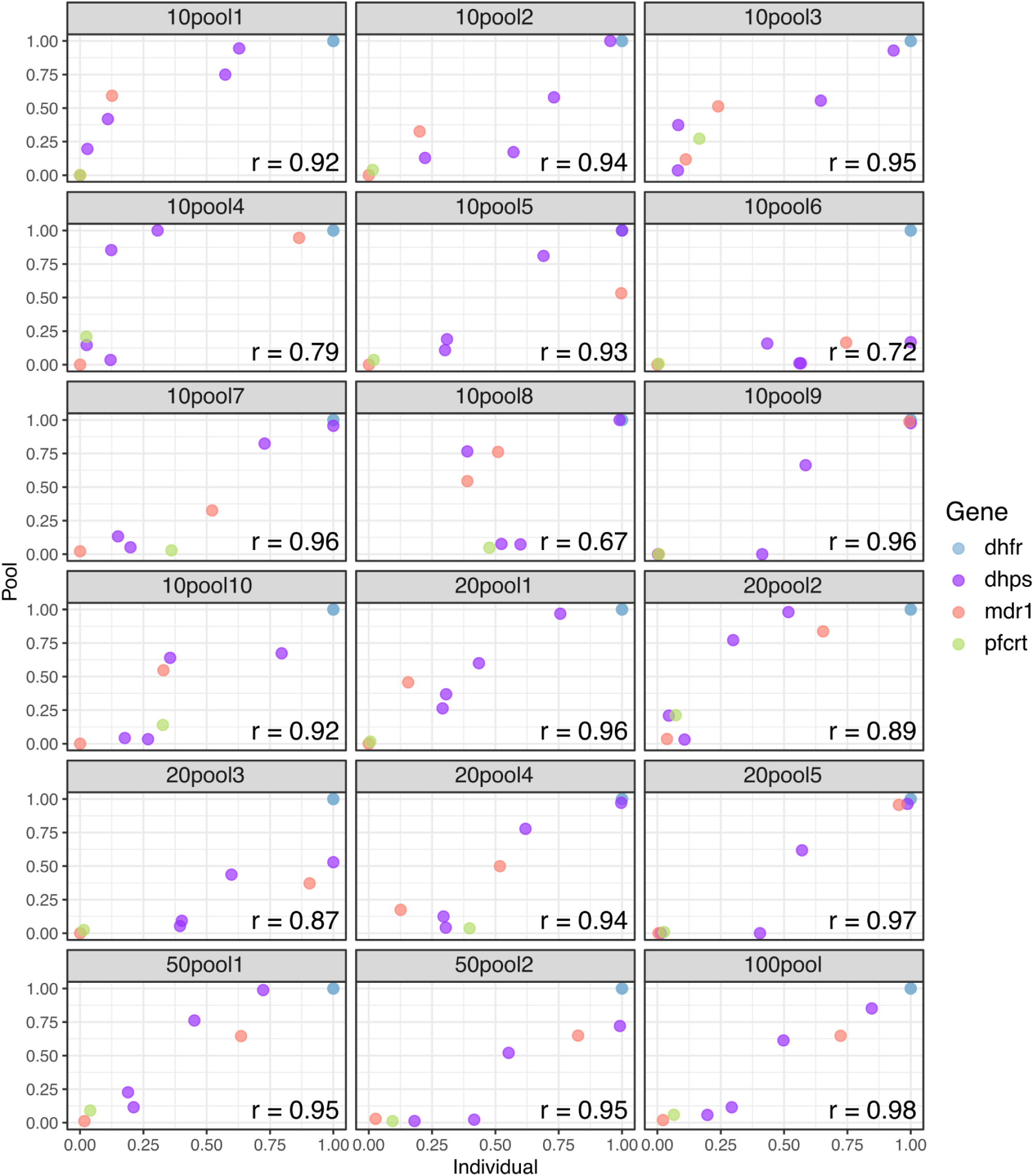
Correlation of antimalarial resistance SNP frequency in pools compared to individual samples. Weighted mean allele frequency for key resistance mutations was estimated across replicates for each pool group and compared with allele frequencies across replicates for the individual samples included in each pool. Allele frequencies in the pool were weighted by parasite density of each individual sample included in the pool. Pearson’s correlation values were high across all pool groups. Each frame is labeled as ##pool#, where ## is the pool size and # is the pool number.

## DISCUSSION

We report the development and validation of a highly multiplexed amplicon deep sequencing tool *Pf*-SMARRT and successful application to field samples from Cameroon. Our goal was to develop a simple assay that was freely available using standard reagents to characterize key antimalarial drug resistance genes and provide basic information on parasite COI, diversity, and relatedness. The assay leverages no custom products and the necessary protocols are available within this publication. *Pf*-SMARRT provided robust data across a range of parasitemias, with most amplicons successfully genotyped at 1 p/µL. The assay was then used to characterize antimalarial resistance profiles in Dschang, Cameroon, with most samples genotyped at all amplicons successfully. The assay was compared to MIPs in a subset of samples and showed an overall similar pattern of parasite population clustering based on PCA but demonstrated a higher rate of calling minor haplotypes, resulting in higher prevalence for drug resistance molecular markers and higher COI estimates. Lastly, we evaluated the assay using pooled deep sequencing to characterize how well it would detect antimalarial resistance SNPs in sample pools of different sizes, showing that increased pool size improves the accuracy of frequency calls but can result in missed low-frequency SNPs in the pool. *Pf*-SMARRT has multiple advantageous characteristics, which make it particularly useful for conducting MMS.

Among samples collected in Dschang, Cameroon, in 2020, we found similar results as other recent molecular studies in other parts of the country. Although no validated or candidate K13 markers were found, one non-synonymous polymorphism was identified, the A578**S** mutation, which is commonly seen at low prevalence in Africa but not associated with resistance. Most parasites contained the wild type **N**86Y MDR1 allele, associated with decreased susceptibility to lumefantrine. The CRT K76**T** mutant allele was found in 19% (95% CI:11.3-26.7%) of samples, which is higher than the few reports of this mutation in samples collected at a similar time (**Table 1**). DHFR mutations remain common and DHPS mutations are variable. The K540**E** mutation remains uncommon, similar to previous reports and as seen in West Africa (Okell et al., 2017). We detected an appreciable number of I431**V** mutants (38%, CI: 28.2-47.0), a mutation that has been found and may be increasing in Ghana, Nigeria and Cameroon (Oguike et al., 2016; Adegbola et al., 2023). In addition, the DHFR A613**S** was found at a prevalence of 31.0% (CI: 21.6-40.1).

When comparing *Pf*-SMARRT to MIPs, a higher number of mixed infections were found with *Pf*-SMARRT than with MIPs (**Table 2**). This resulted in an increased estimated prevalence for most markers (**Table 3**). Overall, *Pf*-SMARRT identified mixed genotypes in 12.5% (81/650) of loci, whereas MIPs found mixed genotypes at 6.4% of loci (40/628). There are likely two reasons for this difference. PCR amplicons may be more efficient than capture-based approaches like MIPs. This is primarily due to the nature of PCR amplification, which enriches specific target regions, ensuring that even minor clones within a sample are more likely to be captured and sequenced. The inherent nature of PCR allows for amplification of low-abundance targets, making it easier to detect mixed genotypes and rare variants. The depth of sequencing is another critical aspect that enhances the performance of *Pf*-SMARRT. Deeper sequencing of a smaller number of loci allows for a more finely articulated view of the genetic diversity within a sample, making it possible to detect low-frequency alleles that may exist in mixed infections.

This increased depth is particularly valuable in malaria research, where understanding the prevalence of minor variants can inform public health strategies and resistance monitoring. While MIPs can achieve significant coverage, they often operate at shallower sequencing depths (10X-50X coverage). This can lead to a loss of sensitivity in detecting low-density infections, limiting their ability to uncover important genetic variants. It is important to note that while MIPs can, in theory, achieve similar sensitivity to *Pf*-SMARRT with increased coverage and depth, the practical implementation of this is often constrained by factors such as cost and technical complexity. *Pf*-SMARRT stands out as a more effective tool for detecting all clones and resistance mutations identified by MIPs while also capturing more minor clones and rare variants. This enhanced sensitivity is vital in regions where malaria transmission declines and efforts shift toward pre-elimination strategies. The ability to detect low-density infections and subtle genetic variations makes *Pf*-SMARRT particularly useful for MMS, enabling researchers and public health officials to make informed decisions in the fight against malaria.

Importantly, MIPs out performed *Pf*-SMARRT for detecting genetically similar clusters of parasites within the population (**Figure 3**). This is not surprising as the MIP panels used genotyped over 480 amplicons across the genome as compared to 24 amplicons with *Pf*-SMARRT. The use of IBD measurement is inherently affected by the number of amplicons included and their distribution across a genome (Guo et al., 2023). *Pf*-SMARRT was not designed specifically for use with IBD but rather to have power to discern clones based upon global heterozygosity of the diversity amplicons. For example, based on the measured heterozygosity of the population in Cameroon (**Table 4**) we would have a 99.9998% chance of determining strains are different (and 99.9995% if Heome H is not successfully genotyped).

Pooling has previously been proposed as a way to cost effectively monitor parasite variation (Taylor et al., 2013, 2015; Juliano et al., 2016; Brazeau et al., 2019) and has worked well for single amplicon deep sequencing. This was the first time we used highly multiplexed amplicon deep sequencing in pooled samples. The results suggest that sensitivity for rare alleles would be a significant limitation for using pooling for monitoring the emergence of new polymorphism (**Figure S13**). However, at higher frequencies, the ability to detect and determine the relative frequency of alleles occuring at moderate to high frequency was good, especially in pools with higher numbers of samples. Thus, the use of a pooling approach may be limited to monitoring known mutations after emergence. Increasing throughput for individual sample sequencing by making the assay protocol straightforward and scalable is necessary for evaluating introduction or emergence of a mutation.

This study, and the assay do have some limitations. First, this version of the assay is missing coverage for several key drug resistance mutations including P574L in K13, I164L in DHFR and D1246Y in MDR1. In addition, the ligation based library preparation is more complicated and time consuming than a PCR-based library preparation. Modifications to resolve these limitations are underway. Despite sequencing in duplicate and using a relatively conservative minor frequency cut off of 1%, we detected three false positive haplotypes in our control experiments, although all false-positives occurred at the lowest parasitemia, a known issue in amplicon sequencing. This represented only 3 of 288 loci genotyped in the 1 parasite/µL controls. In addition, in our mixture experiment, one amplicon (DHPS 437) failed to detect the Dd2 minor variant consistently. It is unclear what happened during this experiment, as no other indication of problems occurred with this amplicon in the other controls or clinical samples. One of the diversity targets (Heome-H) was consistently under-sequenced relative to other amplicons but had sufficient coverage to allow calls in most samples (76/100). Lastly, we did not directly test the assay in low-density infections from Cameroon (a single low density parasitemia of 0.5 p/µL, likely due to poor extraction, successfully genotyped at 24/24 amplicons), thus we do not know how it will perform using such clinical samples, although the mocked blood spots included whole blood spiked with culture grown parasites and, therefore, should be a good representation.

*Pf*-SMARRT represents a valuable tool for conducting MMS, providing detailed genotyping of 24 amplicons in an open source platform. Control experiments using mocked blood spots and known mixtures of different strains provide support that the assay should accurately report within-sample frequency and reliably detect genotypes in low density infections. The assay had a high success rate in samples with what should be RDT-detectable parasitemia from Dschang, Cameroon. It proved sensitive for minority clones in infections and had a low false positivity rate, which occurred only at very low parasitemia. While pooled sequencing had overall good correlations to individual samples in terms of antimalarial resistance frequency, the lowest frequency alleles were commonly missed in the pools. This suggests that pools may not be appropriate for surveillance of rare alleles in a population but could help monitor the frequency of common alleles. As we move toward malaria elimination, the insights provided by *Pf*-SMARRT will be useful for understanding and monitoring the transmission dynamics of *P. falciparum.* Moreover, as more labs and control programs gain access to next generation sequencers in Africa, the use of MMS to monitor malaria populations will only increase.

*Pf*-SMARRT represents a tool that uses easy to access materials and can easily be implemented across any Illumina platform, including the ISeq, to provide robust characterization of antimalarial resistance polymorphisms and parasite relatedness.

## Acknowledgements

We thank the participants of the study. We thank the laboratory and clinical staff of the Dschang Regional Hospital Annex, the St. Vincent Catholic Hospital and the Catholic Hospital in Batsingla for their assistance in sample collection for this study. The following reagent was obtained through BEI Resources, NIAID, NIH: *Plasmodium falciparum*, Strain 3D7, MRA-102, contributed by Daniel J. Carucci. The following reagent was obtained through BEI Resources, NIAID, NIH: Genomic DNA from *Plasmodium falciparum*, Strain 3D7, MRA-102G, contributed by Daniel J. Carucci. The following reagent was obtained through BEI Resources, NIAID, NIH: Genomic DNA from *Plasmodium falciparum*, Strain Dd2, MRA-150G, contributed by David Walliker.

## Ethics Statement

The study was approved by the institutional review board (IRB) of the Cameroon Baptist Convention Health Board (FWA00002077), Protocol IRB2019-40. Written informed consent was administered in French or English, based on participant preference, via an independent translator (who also spoke the local language Yemba). For children, informed consent was obtained from a parent or guardian. Molecular analysis of de-identified samples and data were deemed non-human subjects research by the University of North Carolina IRB.

## Data Availability

Sequencing data are available on the sequence read archive (PRJNA1171111). Metadata are available in Table S3 and upon request to Dr. Innocent M. Ali. Custom scripts are available at https://github.com/bailey-lab/pfsmarrt_cameroon_7-6-24.

## Author contributions

Conception and design: IMA, JAB, JJJ; Data acquisition, analysis and interpretation: JMB, AS, VPKT, IGG, AAF, KW, KN, KT, SW, CBT, IMA, JAB, JJJ; Drafting and revising: JMS, AS, IGG, AAF, IMA, JAB, JJJ; Final approval: JMS, AS, VPKT, IGG, AF, KW, AA, KN, KT, SW, WM, RD, SN, CBT, JBP, IMA, JAB, JJJ; Accountability: JMS, AS, JAB, JJJ

