## Supplemental Material for "Application of a new highly multiplexed amplicon sequencing tool to evaluate *Plasmodium falciparum* antimalarial resistance and relatedness in individual and pooled samples from Dschang, Cameroon"

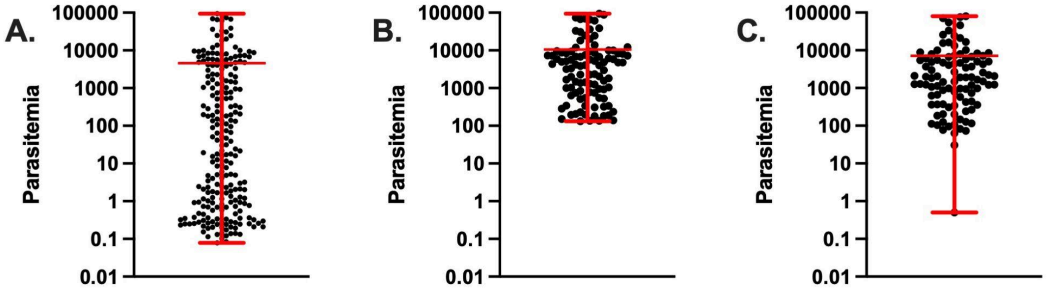

**Figure S1. PCR estimated parasitemias based on real-time PCR.** Panel A shows the distribution of estimated parasitemia in the entire study population. Panel B shows the estimated parasitemias of the 100 selected for this sequencing study. Panel C shows the estimated parasitemia after re-extraction. Red bars show mean and whiskers denote maximum and minimum parasitemia. Parasitemia is shown on a log10 scale.

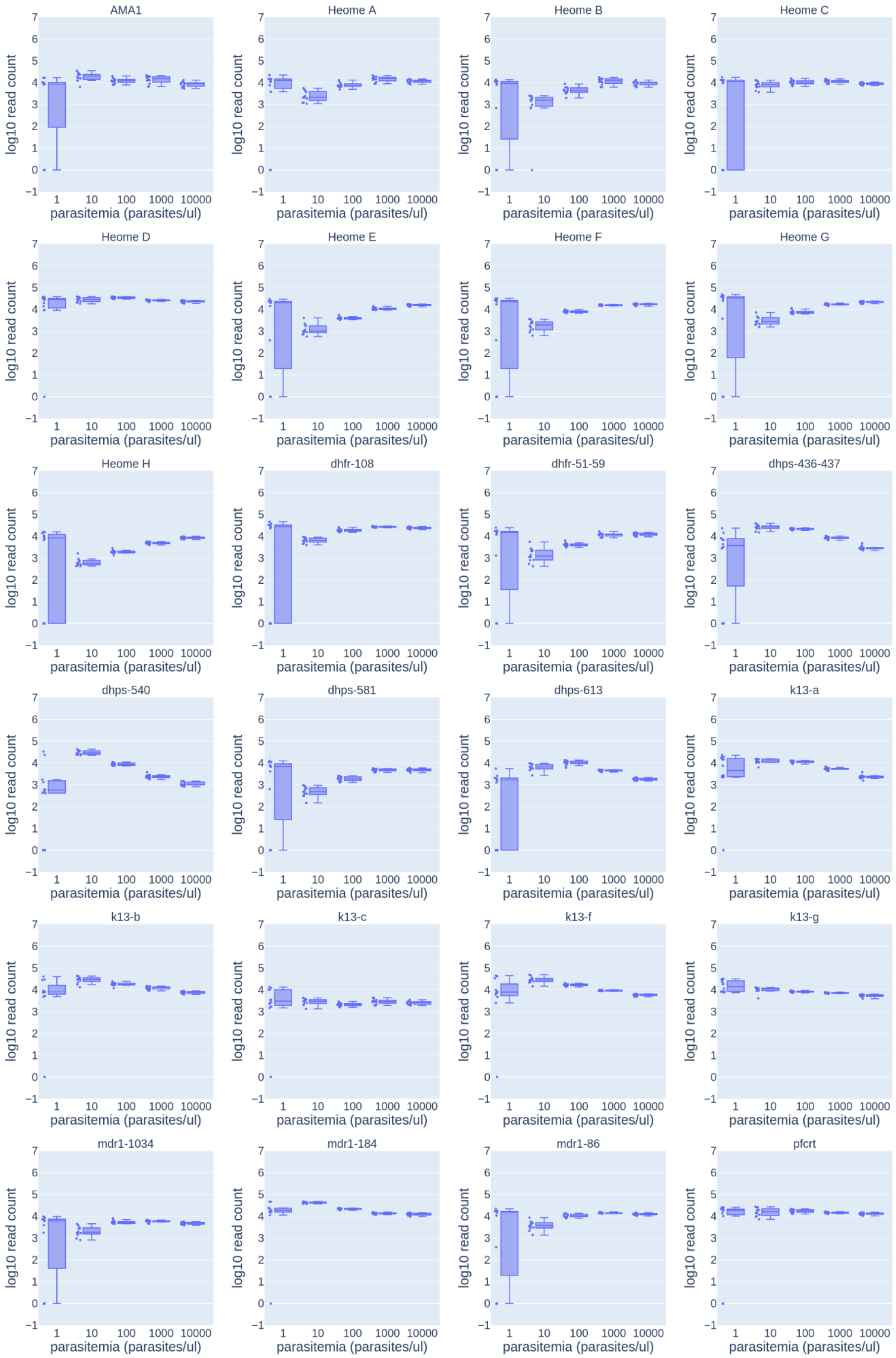

**Figure S2. Sequencing depth for assay development controls.** Plots depict sequencing depth on a log10 scale for 12 samples at each parasitemia. At each parasitemia level, individual points for values are plotted (left) and a box and whiskers plot with maximum and minimum values (right).

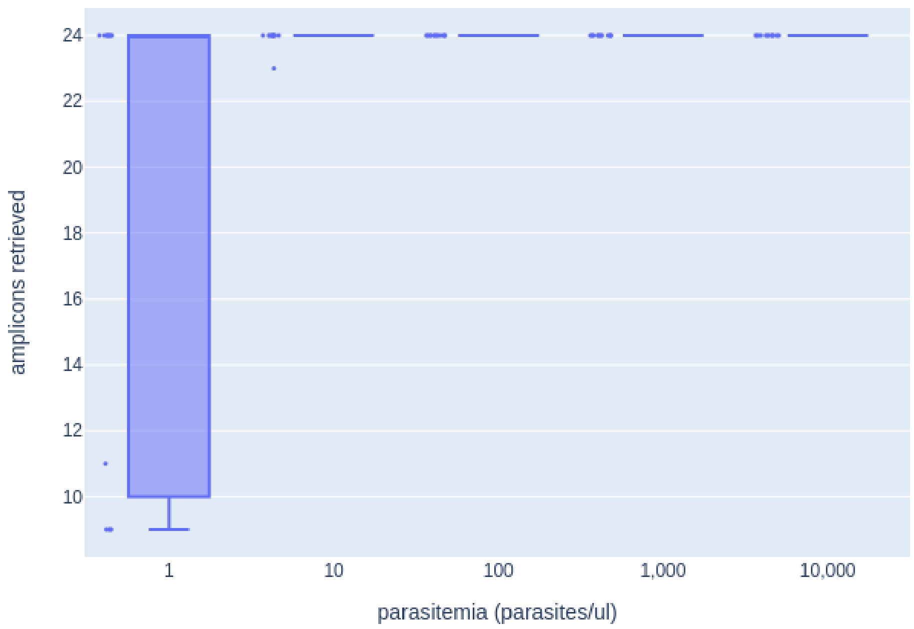

**Figure S3. Number of successfully genotyped amplicons in controls at different parasitemias.** Successfully genotyped amplicons are defined as amplicons that retrieve at least one haplotype within a given replicate. At each parasitemia level, individual points for values are plotted (left) and a box and whiskers plot with maximum and minimum values (right).

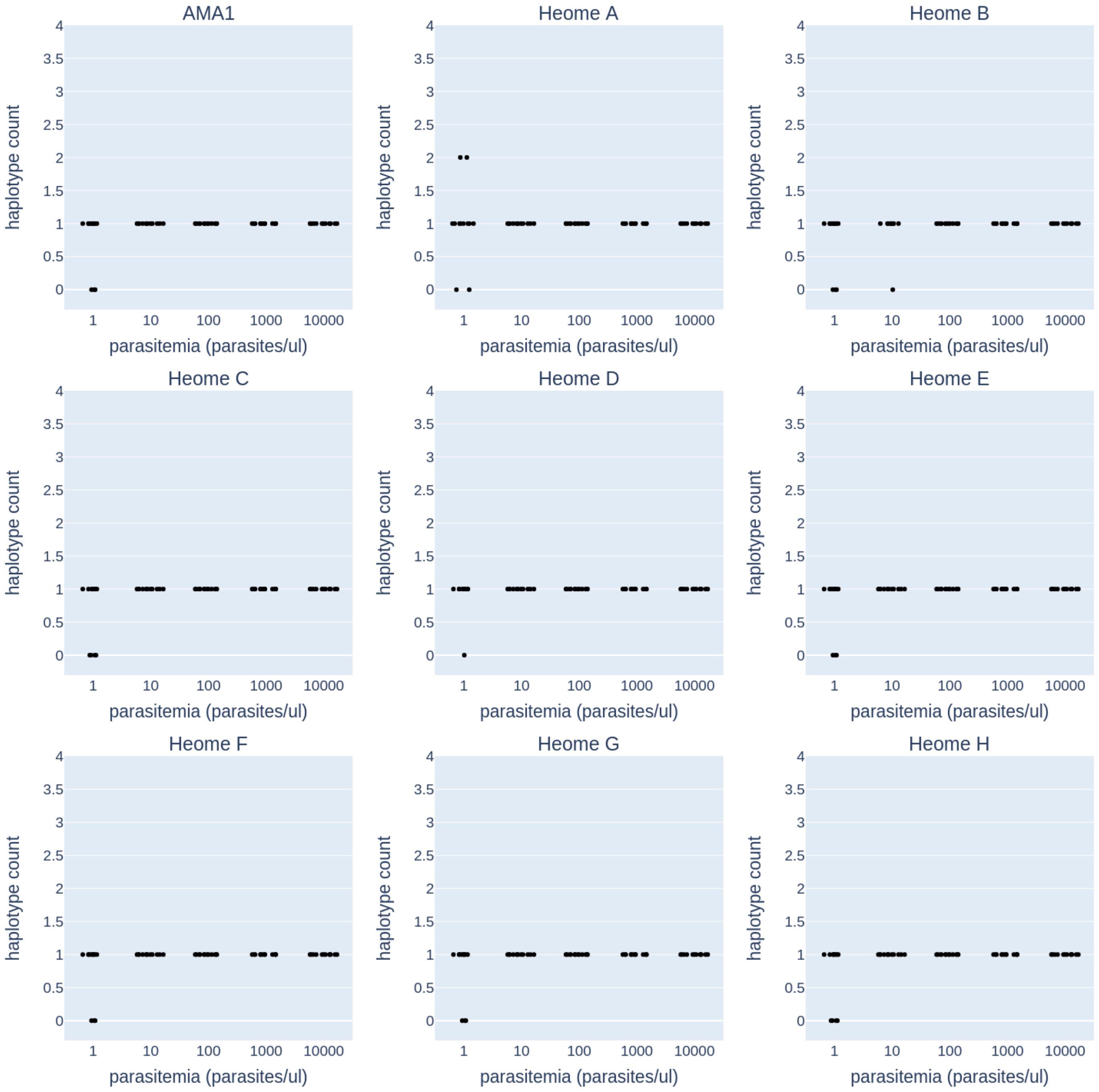

**Figure S4. Haplotype counts in monoclonal control samples at different parasitemias for diversity amplicons**. This figure shows the relationship between parasitemia and the number of haplotypes detected for amplicons that are markers of diversity. There are 12 samples at each parasitemia, and because the samples are monoclonal, the expected number of haplotypes is one.

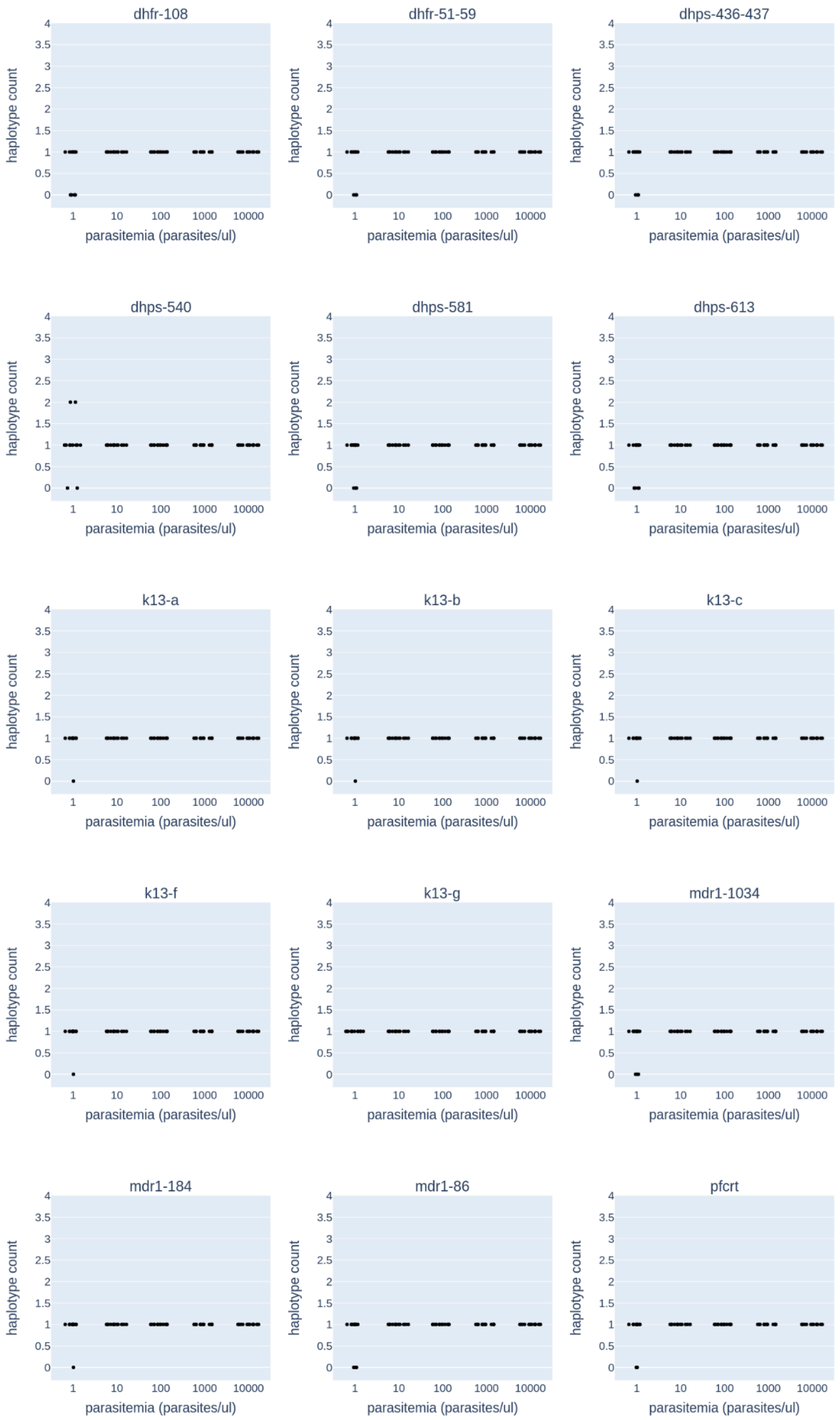

**Figure S5. Haplotype counts in monoclonal control samples at different parasitemias for drug resistance amplicons**. This figure shows the relationship between parasitemia and the number of haplotypes detected for amplicons containing known resistance loci. There are 12 samples at each parasitemia, and because the samples are monoclonal, the expected number of haplotypes is one.

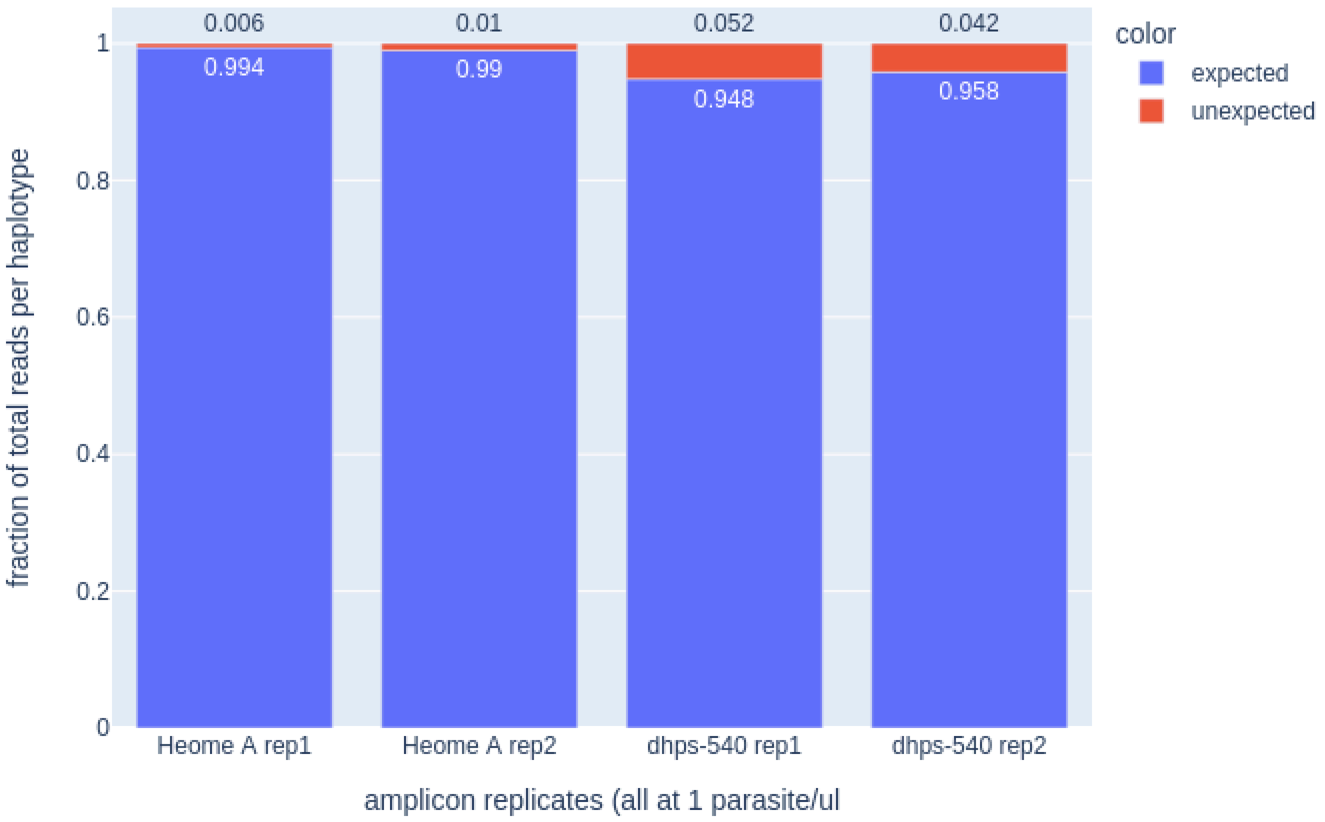

**Figure S6. Control samples with unexpected haplotypes.** A total of 4 replicate/amplicon/parasitemia combinations (of 1,440 genotyped combinations) had unexpected haplotypes detected (red). All of the replicates with unexpected haplotypes occurred at a concentration of 1 parasite per microliter. The expected haplotype was detected in all samples as the majority variant.

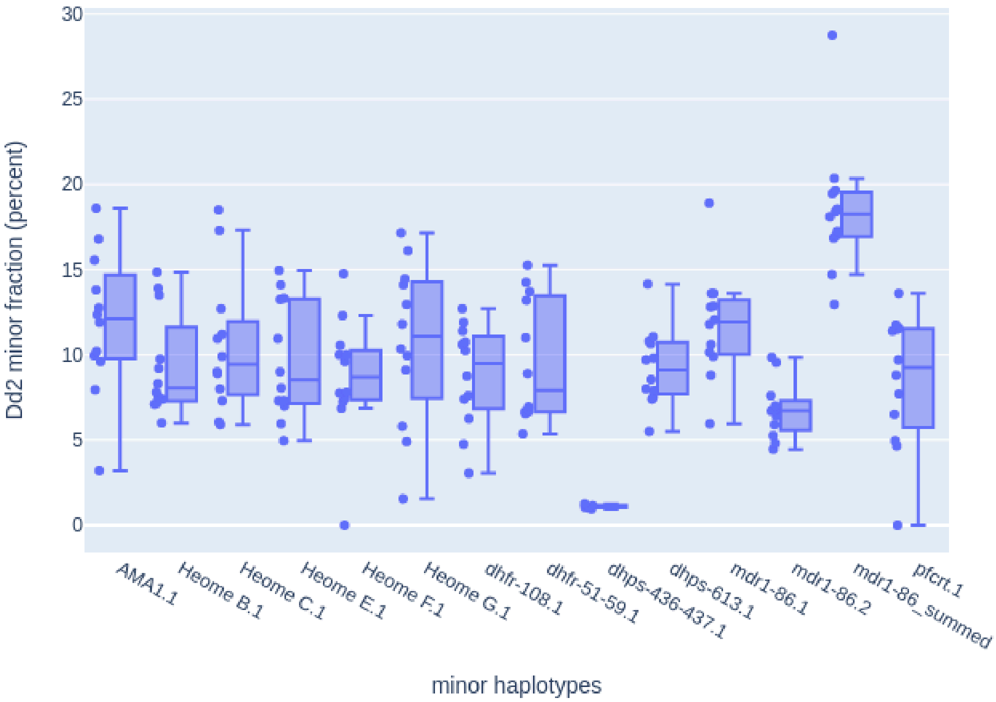

**Figure S7. Within sample allele frequency of known mixture of 3D7 and Dd2 (88:12).** The amplicon specific within sample allele frequency for the Dd2 minor variant are shown for all 12 samples. Only amplicons that are variable between the two strains are shown. The expected allele frequency is 12 percent. Strain Dd2 has an imperfect second copy of MDR1 which alters the allele frequency at these amplicons (both the frequencies of individual copies and the sum of the frequencies of both copies are shown). At each amplcion, individual points for values are plotted (left) and a box and whiskers plot with maximum and minimum values (right).

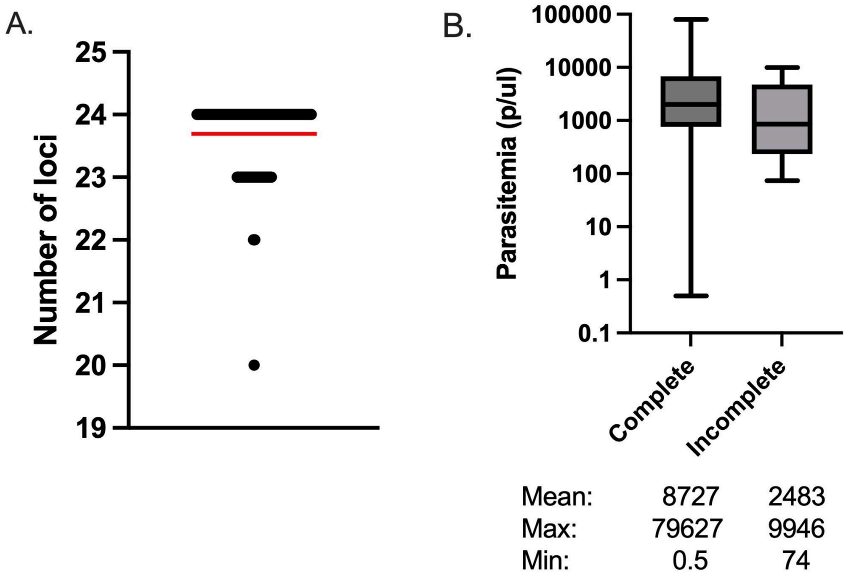

**Figure S8. Number of successfully genotyped amplicons per sample among 100 Cameroonian isolates.** Panel A shows the number of successful amplicons for each sample, where a successful amplicon is one containing at least one haplotype. The red bar is the mean for all 100 samples. Panel B shows the parasitemia distribution of samples with complete (24 amplicons) versus non-complete (<24 amplicons) genotyping ona log10 scale. The line in the box displays the median parasitemia and the whiskers show the minimum and maximum parasitemia (ns, p= 0.07, t-test).

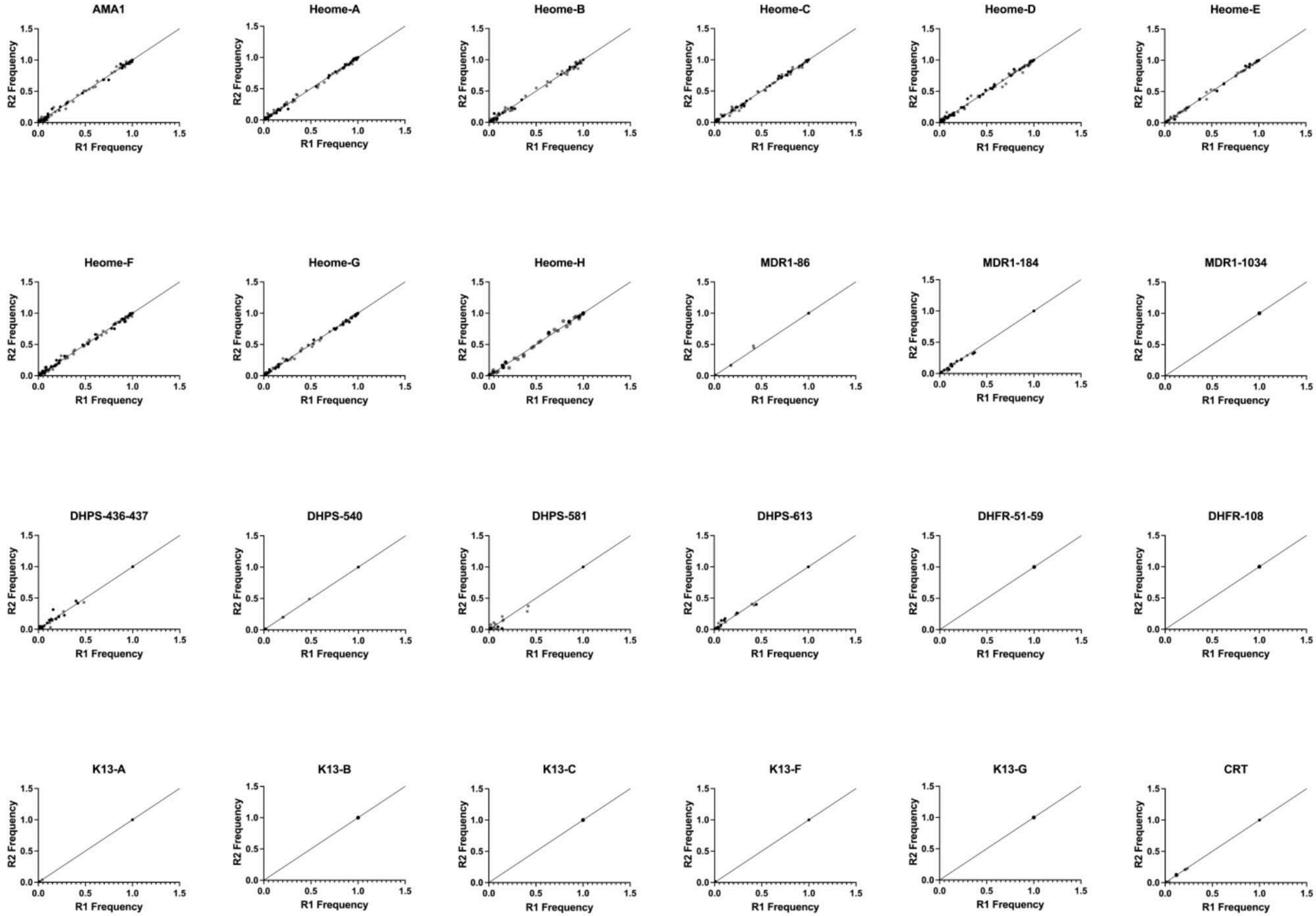
AMA1
Heome-A
Heome-B
Heome-C
Heome-D
Heome-E
Heome-F
Heome-G
Heome-H
MDR1-86
MDR1-184
MDR1-1034
DHPS-436-437
DHPS-540
DHPS-581
DHPS-613
DHFR-51-59
DHFR-108
K13-A
K13-B
K13-C
K13-F
K13-G
CRT
R2 Frequency
R1 Frequency
0.0
0.5
1.0
1.5

**Figure S9. Within sample allele frequency comparison of replicates by amplicon for 100 Cameroonian isolates.** The calculated within sample haplotype frequency (diversity markers) and antimalarial single nucleotide polymorphisms (SNP) are shown for each replicate. Black dots represent samples with two replicates. For samples where multiple haplotypes were detected for resistance genes, the lowest frequency haplotype was reported.

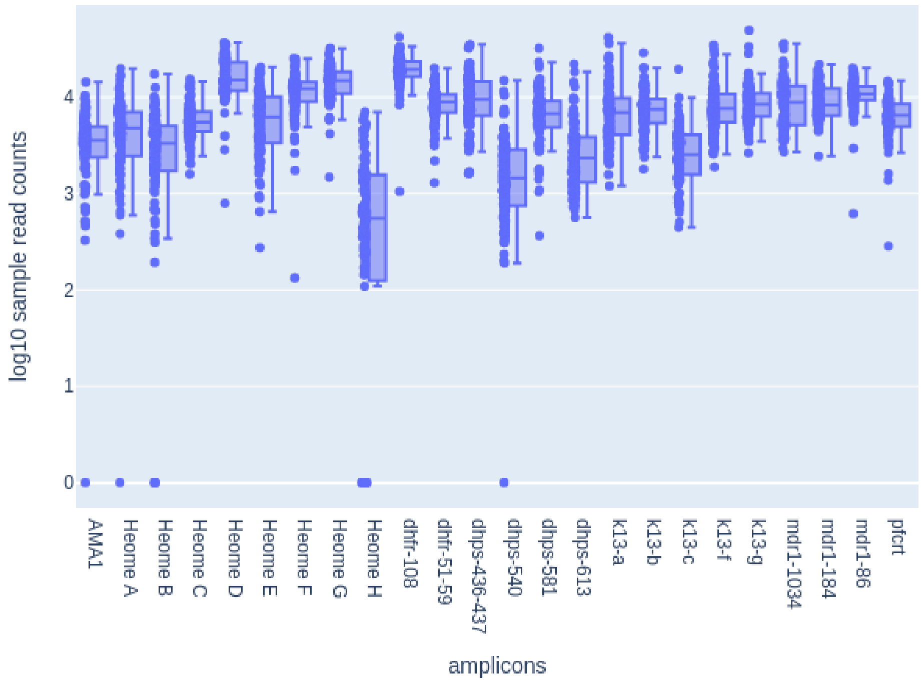

**Figure S10. Read depth at each amplicon for 100 Cameroonian isolates.** Each amplicon contains 100 Cameroonian isolates. Each isolate contains two replicates which are averaged to yield a single amplicon read count per isolate, illustrated by 100 points per amplicon. At each amplicon, individual points for values are plotted (left) and a box and whiskers plot with maximum and minimum values (right).

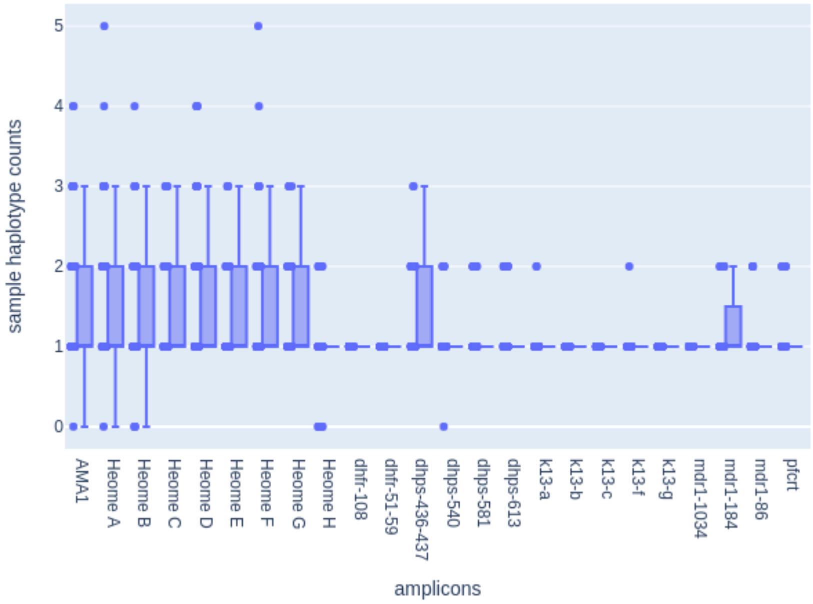

**Figure S11. Complexity of infection (COI) at each amplicon for 100 Cameroonian isolates.** The COI for each amplicon from the samples is shown. At each amplicon, individual points for values are plotted (left) and a box and whiskers plot with maximum and minimum values (right).

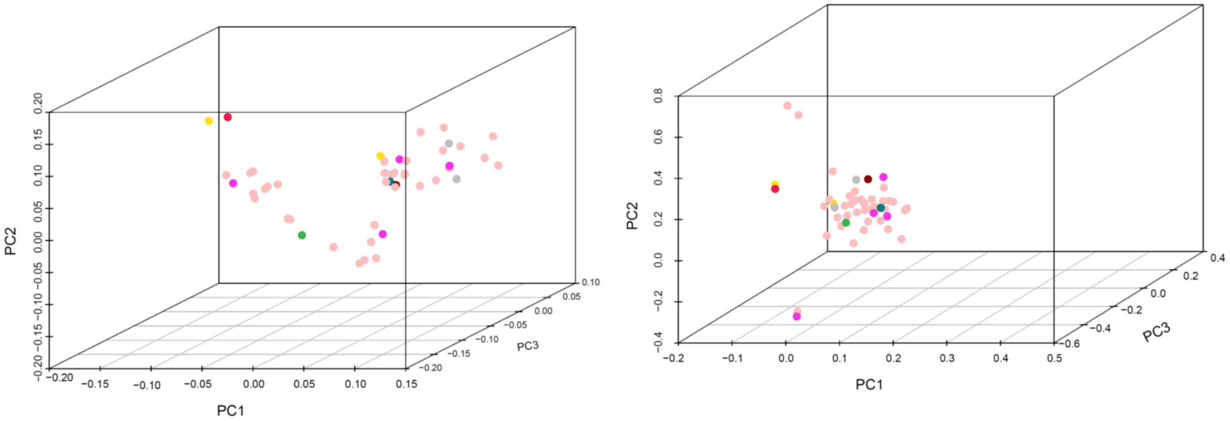

**Figure S12. Principal Component Analysis of 50 Cameroonian Samples using *Pf*-SMARRT (A) and Molecular Inversion Probes (MIP)**

A

B

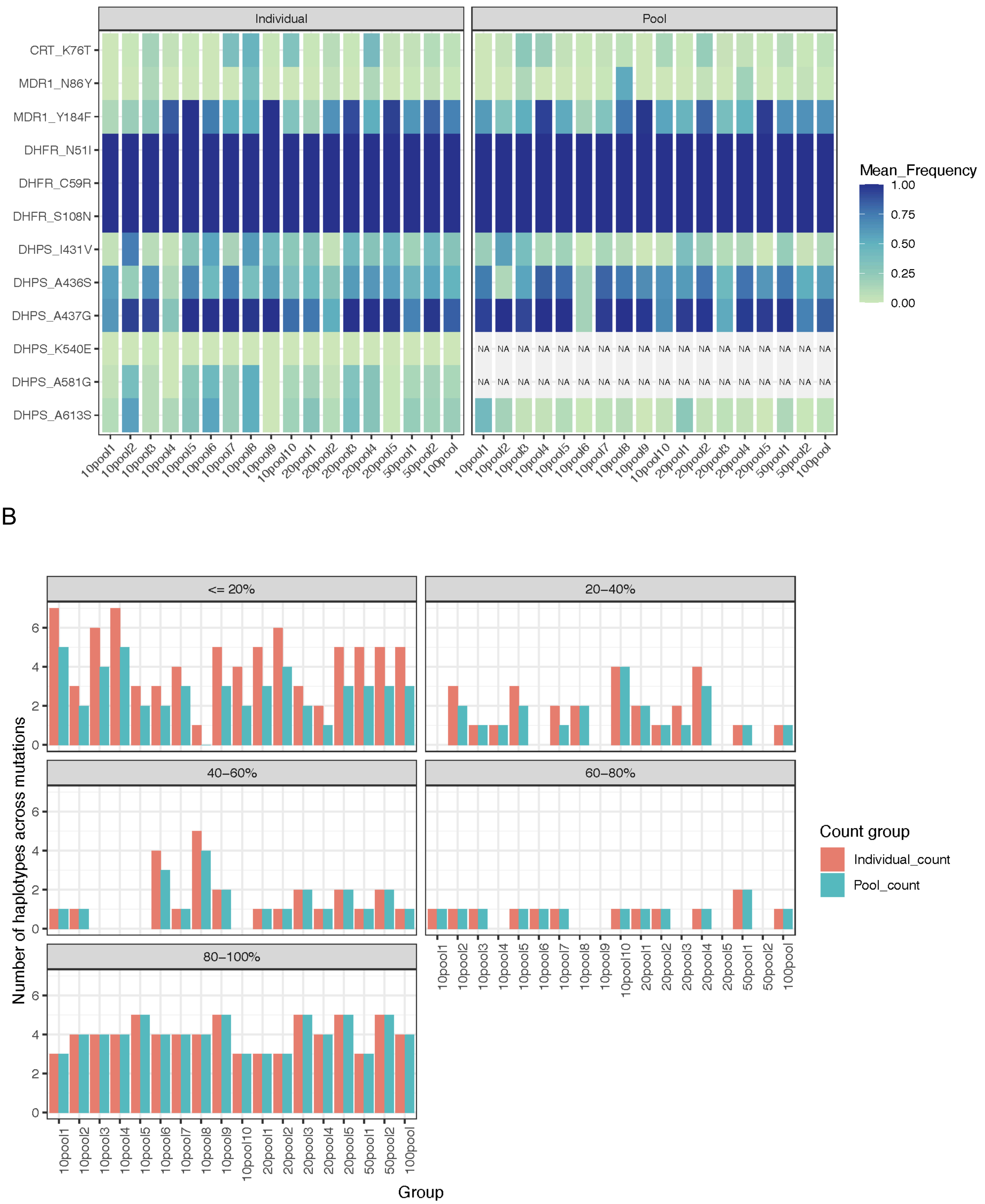

**Figure S13. Allele frequency in pooled samples.** Weighted mean allele frequency for key resistance mutations shows consistency between pooled and individual samples. Exceptions occur for highly variable and low-frequency variants such as mutations K540E and A581G with frequencies varying from 0.0-0.7% and 0.1-51.5%, respectively, that are not found in pooled samples (A). Lower consistency between the number of total mutations found in pooled versus individual samples can be seen in variants that occur with a frequency lower than 40%, whereas more frequent mutations are successfully identified in both pooled and individual samples (B).

**Table S1. Comparison of population characteristics**

| | Not sequenced (n=130) | Sequenced (n=100) | Total (N=230) | P-value |
|---|---|---|---|---|
| **Age (years)** | 28.0 (21.0, 45.0) | 25.0 (19.0, 35.0) | 26.0 (20.0, 40.0) | 0.28 |
| **Sex?** | | | | 0.09 |
| Male | 53 (41%) | 52 (52%) | 105 (46%) | |
| Female | 77 (59%) | 48 (48%) | 125 (54%) | |
| **Education** | | | | 0.35 |
| Primary | 55 (42%) | 33 (33%) | 88 (38%) | |
| Secondary | 20 (15%) | 17 (17%) | 37 (16%) | |
| University | 55 (42%) | 50 (50%) | 105 (46%) | |
| **Occupation** | | | | 0.60 |
| Self-employed | 36 (72%) | 23 (82%) | 59 (76%) | |
| Employer | 5 (10%) | 2 (7%) | 7 (9%) | |
| Not working | 9 (18%) | 3 (11%) | 12 (15%) | |
| **Headache?** | | | | 0.72 |
| No | 90 (69%) | 67 (67%) | 157 (68%) | |
| Yes | 40 (31%) | 33 (33%) | 73 (32%) | |
| **General fatigue?** | | | | 0.56 |
| No | 93 (72%) | 68 (68%) | 161 (70%) | |
| Yes | 37 (28%) | 32 (32%) | 69 (30%) | |
| **Vomiting?** | | | | 0.59 |
| No | 122 (94%) | 92 (92%) | 214 (93%) | |
| Yes | 8 (6%) | 8 (8%) | 16 (7%) | |
| **Cough?** | | | | 0.59 |
| No | 122 (94%) | 92 (92%) | 214 (93%) | |
| Yes | 8 (6%) | 8 (8%) | 16 (7%) | |
| **Abdominal pain?** | | | | 0.19 |
| No | 90 (69%) | 61 (61%) | 151 (66%) | |
| Yes | 40 (31%) | 39 (39%) | 79 (34%) | |
| **Gastric pain?** | | | | 0.21 |
| No | 128 (98%) | 100 (100%) | 228 (99%) | |
| Yes | 2 (2%) | | 2 (1%) | |
| **Other symptoms?** | | | | 0.47 |
| No | 122 (94%) | 96 (96%) | 218 (95%) | |
| Yes | 8 (6%) | 4 (4%) | 12 (5%) | |
| **Slept under a bednet in the last 24 hours?** | | | | 0.11 |
| No | 55 (42%) | 32 (32%) | 87 (38%) | |
| Yes | 75 (58%) | 68 (68%) | 143 (62%) | |
| **Nights in the last 7 slept under a bednet?** | 7.0 (0.0, 7.0) | 7.0 (0.0, 7.0) | 7.0 (0.0, 7.0) | 0.35 |
| **Number of people in the household?** | 2.5 (2.0, 5.0) | 3.0 (2.0, 5.0) | 3.0 (2.0, 5.0) | 0.42 |
| **Number of bednets in the household?** | 1.0 (0.0, 2.0) | 1.0 (0.0, 2.0) | 1.0 (0.0, 2.0) | 0.73 |
| **Water within a 2 minutes walk of the household?** | | | | 0.17 |
| No | 102 (78%) | 79 (79%) | 181 (79%) | |
| Yes | 21 (16%) | 20 (20%) | 41 (18%) | |
| Don't know | 7 (5%) | 1 (1%) | 8 (3%) | 0.17 |
| **Type of water source?** | | | | 0.28 |
| Stream | 3 (2%) | 6 (6%) | 9 (4%) | |
| Pond/lake | 9 (7%) | 7 (7%) | 16 (7%) | |
| Swamp/marsh | 9 (7%) | 7 (7%) | 16 (7%) | |
| None | 102 (78%) | 79 (79%) | 181 (79%) | |
| Don't know | 7 (5%) | 1 (1%) | 8 (3%) | |
| **Traveled in the last 3 months?** | | | | 0.34 |
| No | 78 (84%) | 53 (78%) | 131 (81%) | |

| | | | | |
|---|---|---|---|---|
| Yes | 15 (16%) | 15 (22%) | 30 (19%) | |
| **Is a member of the household working in the forest?** | | | | 0.85 |
| No | 126 (99%) | 97 (99%) | 223 (99%) | |
| Yes | 1 (1%) | 1 (1%) | 2 (1%) | |
| **Animals in the house?** | | | | 0.02 |
| No | 120 (94%) | 84 (86%) | 204 (91%) | |
| Yes | 7 (6%) | 14 (14%) | 21 (9%) | |
| **Pregnant?** | | | | 0.96 |
| No | 122 (96%) | 94 (96%) | 216 (96%) | |
| Yes | 5 (4%) | 4 (4%) | 9 (4%) | |
| **Taken antimalarials in the last 14 days?** | | | | 0.63 |
| No | 93 (73%) | 70 (72%) | 163 (73%) | |
| Yes | 28 (22%) | 24 (25%) | 52 (23%) | |
| Don't know | 4 (3%) | 3 (3%) | 7 (3%) | |
| Refused | 2 (2%) | | 2 (1%) | |
| **Which antimalarials?** | | | | 0.51 |
| Quinine | 5 (15%) | 4 (15%) | 9 (15%) | |
| ACT | 16 (47%) | 11 (41%) | 27 (44%) | |
| Don't know | 11 (32%) | 12 (44%) | 23 (38%) | |
| Refused | 2 (6%) | | 2 (3%) | |
| **Source of antimalarials?** | | | | 0.56 |
| Drug seller | 6 (22%) | 8 (35%) | 14 (28%) | |
| Clinic | 14 (52%) | 11 (48%) | 25 (50%) | |
| Leftover | 7 (26%) | 4 (17%) | 11 (22%) | |
| **Lifelong malaria exposure?** | | | | 0.34 |
| No | 9 (7%) | 4 (4%) | 13 (6%) | |
| Yes | 118 (93%) | 94 (96%) | 212 (94%) | |
| **Number of malaria exposures over lifetime?** | 7.0 (3.0, 7.0) | 7.0 (6.5, 7.0) | 7.0 (5.0, 7.0) | 0.51 |
| **Number of malaria exposures in the last year?** | 0.0 (0.0, 1.0) | 0.0 (0.0, 1.0) | 0.0 (0.0, 1.0) | 0.62 |
| **Has someone in the household been treated for malaria in the last year?** | | | | 0.77 |
| No | 14 (11%) | 12 (12%) | 26 (12%) | |
| Yes | 104 (82%) | 76 (78%) | 180 (80%) | |
| Don't know | 8 (6%) | 8 (8%) | 16 (7%) | |
| Refused | 1 (1%) | 2 (2%) | 3 (1%) | |

P-value comparisons across groups for categorical variables are based on chi-square test of homogeneity; p-values for continuous variables are based on Kruskal-Wallis test for median.

**Table S2. Within sample allele frequency of known mixture of 3D7 and Dd2 (88:12)^**

| | AMA1 | Heome B | Heome C | Heome E | Heome F | Heome G | DHFR 51/59 | DHFR 108 | DHPS 436/437* | DHPS 613 | MDR1 86 (HAP 1) | MDR1 86 (HAP2) | MDR1 86 (TOTAL) | CRT |
|---|---|---|---|---|---|---|---|---|---|---|---|---|---|---|
| Average | 0.119 | 0.094 | 0.105 | 0.097 | 0.087 | 0.107 | 0.096 | 0.088 | 0.011 | 0.093 | 0.118 | 0.068 | 0.185 | 0.085 |
| STDEV | 0.042 | 0.030 | 0.040 | 0.035 | 0.036 | 0.047 | 0.036 | 0.030 | 0.001 | 0.023 | 0.032 | 0.017 | 0.038 | 0.039 |

*: when detected (5/12 reps)

^: Not all loci are variable between the strains. Those with no polymorphisms are not reported.

**Table S3. Individual characteristics and sequencing results for 100 participants sequenced with *Pf*-SMARRT**

See the uploaded Excel file.

- Tab 1: Individual metadata and averaged frequencies for antimalarial resistance polymorphisms and diversity amplicons in each individual
- Tab 2: Data dictionary for metadata
- Tab 3: Underlying sequencing depth (count) data for sequencing replicates for each individual used to calculate within sample allele frequencies

**Table S4. Haplotypes detected for AMA in 100 Cameroonian participants**

| Haplotype Name | Haplotype Sequence |
|---|---|
| ama1.00 | ATTTGGTAAAGGTATAATTATTGAGAATTCAAAAACTACTTTTTTAACACCGGTAGCTACGGAAAATCAAGATTTAAAAGATGGAGGTTTTGCTTTTCCTCCAACAAATCCTCCTATGTCACCAATGACATTAAATGGTATGAGAGATTTATATAAAAATAATGAATATGTAAAAAATTTAGATGAATTGACTTTA |
| ama1.01 | ATTTGGTAAAGGTATAATTATTGAGAATTCAAATACTACTTTTTTAACACCGGTAGCTACGGAAAATCAAGATTTAAAAGATGGAGGTTTTGCTTTTCCTCCAACAAATCCTCTTATATCACCAATGACATTAGATCATATGAGAGATTCTTATAAAAATAATGAATATGTAAAAAATTTAGATGAATTGACTTTA |
| ama1.02 | ATTTGGTAAAGGTATAATTATTGAGAATTCAAATACTACTTTTTTAAAACCGGTAGCTACGGGAAATCAAGATTTAAAAGATGGAGGTTTTGCTTTTCCTCCAACAAATCCTCTTATATCACCAATGACATTAAATGGTATGAGAGATTTTTATAAAAATAATGAATATGTAAAAAATTTAGATGAATTGACTTTA |
| ama1.03 | ATTTGGTAAAGGTATAATTATTGAGAATTCAAATACTACTTTTTTAACACCGGTAGCTACGGAAAATCAAGATTTAAAAGATGGAGGTTTTGCTTTTCCTCCAACAAAACCTCTTATGTCACCAATGACATTAGATCAAATGAGACATTTTTATAAAGATAATAAATATGTAAAAAATTTAGATGAATTGACTTTA |
| ama1.04 | ATTTGGTAAAGGTATAATTATTGAGAATTCAAATACTACTTTTTTAACACCGGTAGCTACGGGAAATCAATATTTAAAAGATGGAGGTTTTGCTTTTCCTCCAACAGAACCTCATATGTCACCAATGACATTAGATGAAATGAGACATTTTTATAAAGATAATAAATATGTAAAAAATTTAGATGAATTGACTTTA |
| ama1.05 | ATTTGGTAAAGGTATAATTATTGAGAATTCAAATACTACTTTTTTAAAACCGGTAGCTACGGGAAATCAAGATTTAAAAGATGGAGGTTTTGCTTTTCCTCCAACAGAACCTCTTATATCACCAATGACATTAGATGATATGAGAGATTTTTATAAAAATAATGAATATGTAAAAAATTTAGATGAATTGACTTTA |
| ama1.06 | ATTTGGTAAAGGTATAATTATTGAGAATTCAAATACTACTTTTTTAACACCGGTAGCTACGGAAAATCAAGATTTAAAAGATGGAGGTTTTGCTTTTCCTCCAACAGAACCTCTTATGTCACCAATGACATTAGATCAAATGAGACATTTTTATAAAGATAATAAATATGTAAAAAATTTAGATGAATTGACTTTA |
| ama1.07 | ATTTGGTAAAGGTATAATTATTGAGAATTCAAATACTACTTTTTTAACACCGGTAGCTACGGAAAATCAAGATTTAAAAGATGGAGGTTTTGCTTTTCCTCCAACAAAACCTCTTATGTCACCAATGACATTAGATCAAATGAGAGATTTTTATAAAAATAATGAATATGTAAAAAATTTAGATGAATTGACTTTA |
| ama1.08 | ATTTGGTAAAGGTATAATTATTGAGAATTCAAATACTACTTTTTTAACACCGGTAGCTACGGAAAATCAAGATTTAAAAGATGGAGGTTTTGCTTTTCCTCCAACAAAACCTCTTATGTCACCAATGACATTAGATGAAATGAGACATTTTTATAAAGATAATAAATATGTAAAAAATTTAGATGAATTGACTTTA |
| ama1.09 | ATTTGGTAAAGGTATAATTATTGAGAATTCAAATACTACTTTTTTAAAACCGGTAGCTACGGGAAATCAAGATTTAAAAGATGGAGGTTTTGCTTTTCCTCCAACAGAACCTCTTATATCACCAATGACATTAAATGGTATGAGAGATTTTTATAAAAATAATGAATATGTAAAAAATTTAGATGAATTGACTTTA |
| ama1.10 | ATTTGGTAAAGGTATAATTATTGAGAATTCAAATACTACTTTTTTAACACCGGTAGCTACGGGAAATCAATATTTAAAAGATGGAGGTTTTGCTTTTCCTCCAACAGAACCTCTTATGTCACCAATGACATTAGATGAAATGAGACATTTTTATAAAGATAATAAATATGTAAAAAATTTAGATGAATTGACTTTA |
| ama1.11 | ATTTGGTAAAGGTATAATTATTGAGAATTCAAATACTACTTTTTTAACACCGGTAGCTACGGGAAAACAAGATTTAAAAGATGGAGGTTTTGCTTTTCCTCCAACAAATCCTCTTATATCACCAATGACATTAAATGGTATGAAAGATTTTTATAAAGATAATGAAGATGTAAAAAATTTAGATGAATTGACTTTA |
| ama1.12 | ATTTGGTAAAGGTATAATTATTGAGAATTCAAAAACTACTTTTTTAACACCGGTAGCTACGGAAAATCAAGATTTAAAAGATGGAGGTTTTGCTTTTCCTCCAACAAAACCTCTTATGTCACCAATGACATTAGATGATATGAGAGATCTTTATAAAAATAATGAATATGTAAAAAATTTAGATGAATTGACTTTA |
| ama1.13 | ATTTGGTAAAGGTATAATTATTGAGAATTCAAAAACTACTTTTTTAACACCGGTAGCTACGGAAAATCAAGATTTAAAAGATGGAGGTTTTGCTTTTCCTCCAACAGAACCTCTTATGTCACCAATGACATTAGATGATATGAGACGTTTTTATAAAGATAATGAATATGTAAAAAATTTAGATGAATTGACTTTA |
| ama1.14 | ATTTGGTAAAGGTATAATTATTGAGAATTCAAATACTACTTTTTTAACACCGGTAGCTACGGGAAAACAAGATTTAAAAGATGGAGGTTTTGCTTTTCCTCCAACAAATCCTCTTATATCACCAATGACATTAGATCATATGAGAGATTTTTATAAAAAAAATGAATATGTAAAAAATTTAGATGAATTGACTTTA |
| ama1.15 | ATTTGGTAAAGGTATAATTATTGAGAATTCAAATACTACTTTTTTAACACCGGTAGCTACGGGAAATCAAGATTTAAAAGATGGAGGTTTTGCTTTTCCTCCAACAAAACCTCTTATGTCACCAATGACATTAGATGATATGAGACTTTTGTATAAAGATAATGAAGATGTAAAAAATTTAGATGAATTGACTTTA |
| ama1.16 | ATTTGGTAAAGGTATAATTATTGAGAATTCAAAAACTACTTTTTTAACACCGGTAGCTACGGAAAATCAAGATTTAAAAGATGGAGGTTTTGCTTTTCCTCCAACAAAACCTCTTATGTCACCAATGACATTAGATGAAATGAGACATTTTTATAAAGATAATAAATATGTAAAAAATTTAGATGAATTGACTTTA |
| ama1.17 | ATTTGGTAAAGGTATAATTATTGAGAATTCAAATACTACTTTTTTAACACCGGTAGCTACGGAAAATCAAGATTTAAAAGATGGAGGTTTTGCTTTTCCTCCAACAGAACCTCTTATGTCACCAATGACATTAGATCGTATGAGAGATTTTTATAAAAATAATGAAGATGTAAAAAATTTAGATGAATTGACTTTA |

| | |
|---|---|
| ama1.18 | ATTTGGTAAAGGTATAATTATTGAGAATTCAAAAACTACTTTTTTAACACCGGTAGCTACGGAAAATCAAGATTTAAAAGATGGAGGTTTTGCTTTTCCTCCAACAGAACCTCTTATATCACCAATGACATTAGATCAAATGAGACATCTTTATAAAGATAATGAATATGTAAAAAATTTAGATGAATTGACTTTA |
| ama1.19 | ATTTGGTAAAGGTATAATTATTGAGAATTCAAATACTACTTTTTTAACACCGGTAGCTACGGAAAATCAAGATTTAAAAGATGGAGGTTTTGCTTTTCCTCCAACAAATCCTCCTATGTCACCAATGACATTAGATGATATGAGACTTTTGTATAAAGATAATGAAGATGTAAAAAATTTAGATGAATTGACTTTA |
| ama1.20 | ATTTGGTAAAGGTATAATTATTGAGAATTCAAATACTACTTTTTTAACACCGGTAGCTACGGAAAATCAAGATTTAAAAGATGGAGGTTTTGCTTTTCCTCCAACAAATCCTCCTATGTCACCAATGACATTAGATCAAATGAGACATTTTTATAAAGATAATAAATATGTAAAAAATTTAGATGAATTGACTTTA |
| ama1.21 | ATTTGGTAAAGGTATAATTATTGAGAATTCAAAAACTACTTTTTTAACACCGGTAGCTACGGAAAATCAAGATTTAAAAGATGGAGGTTTTGCTTTTCCTCCAACAAAACCTCTTATGTCACCAATGACATTAGATGATATGAGAGATTTTTATAAAGATAATGAATATGTAAAAAATTTAGATGAATTGACTTTA |
| ama1.22 | ATTTGGTAAAGGTATAATTATTGAGAATTCAAATACTACTTTTTTAACACCGGTAGCTACGGAAAATCAAGATTTAAAAGATGGAGGTTTTGCTTTTCCTCCAACAAAACCTCATATGTCACCAATGACATTAGATCAAATGAGACATTTTTATAAAGATAATAAATATGTAAAAAATTTAGATGAATTGACTTTA |
| ama1.23 | ATTTGGTAAAGGTATAATTATTGAGAATTCAAATACTACTTTTTTAACACCGGTAGCTACGGAAAATCAAGATTTAAAAGATGGAGGTTTTGCTTTTCCTCCAACAAAACCTCTTATGTCACCAATGACATTAGATCAAATGAGACATTTTTATAAAGATAATGAAGATGTAAAAAATTTAGATGAATTGACTTTA |
| ama1.24 | ATTTGGTAAAGGTATAATTATTGAGAATTCAAATACTACTTTTTTAAAACCGGTAGCTACGGGAAATCAAGATTTAAAAGATGGAGGTTTTGCTTTTCCTCCAACAAATCCTCTTATATCACCAATGACATTAAATGGTATGAAAGATTTTTATAAAGATAATGAAGATGTAAAAAATTTAGATGAATTGACTTTA |
| ama1.25 | ATTTGGTAAAGGTATAATTATTGAGAATTCAAATACTACTTTTTTAAAACCGGTAGCTACGGGAAATCAAGATTTAAAAGATGGAGGTTTTGCTTTTCCTCCAACAAATCCTCTTATATCACCAATGACATTAAATGGTATGAGAGATTTATATAAAAATAATGAAGATGTAAAAAATTTAGATGAATTGACTTTA |
| ama1.26 | ATTTGGTAAAGGTATAATTATTGAGAATTCAAAAACTACTTTTTTAACACCGGTAGCTACGGAAAATCAAGATTTAAAAGATGGAGGTTTTGCTTTTCCTCCAACAAAACCTCTTATGTCACCAATGACATTAGATGATATGAGACTTTTGTATAAAGATAATGAAGATGTAAAAAATTTAGATGAATTGACTTTA |
| ama1.27 | ATTTGGTAAAGGTATAATTATTGAGAATTCAAATACTACTTTTTTAACACCGGTAGCTACGGAAAATCAAGATTTAAAAGATGGAGGTTTTGCTTTTCCTCCAACAAAACCTCTTATGTCACCAATGACATTAGATCAAATGAGACATTTTTATAGAGATAATAAATATGTAAAAAATTTAGATGAATTGACTTTA |
| ama1.28 | ATTTGGTAAAGGTATAATTATTGAGAATTCAAAAACTACTTTTTTAACACCGGTAGCTACGGAAAATCAAGATTTAAAAGATGGAGGTTTTGCTTTTCCTCCAACAGAACCTCTTATATCACCAATGACATTAGATGATATGAGAGATTCTTATAAAGATAATGAATATGTAAAAAATTTAGATGAATTGACTTTA |
| ama1.29 | ATTTGGTAAAGGTATAATTATTGAGAATTCAAATACTACTTTTTTAAAACCGGTAGCTACGGGAAATCAAGATTTAAAAGATGGAGGTTTTGCTTTTCCTCCAACAAATCCTCTTATATCACCAATGACATTAGATCAAATGAGACATTTTTATAAAGATAATGAAGATGTAAAAAATTTAGATGAATTGACTTTA |
| ama1.30 | ATTTGGTAAAGGTATAATTATTGAGAATTCAAAAACTACTTTTTTAACACCGGTAGCTACGGAAAATCAAGATTTAAAAGATGGAGGTTTTGCTTTTCCTCCAACAAATCCTCCTATGTCACCAATGACATTAGATGATATGAGACTTTTGTATAAAGATAATGAAGATGTAAAAAATTTAGATGAATTGACTTTA |
| ama1.31 | ATTTGGTAAAGGTATAATTATTGAGAATTCAAATACTACTTTTTTAAAACCGGTAGCTACGGAAAATCAAGATTTAAAAGATGGAGGTTTTGCTTTTCCTCCAACAAATCCTCCTATGTCACCAATGACATTAGATCAAATGAGACATTTTTATAAAGATAATAAATATGTAAAAAATTTAGATGAATTGACTTTA |
| ama1.32 | ATTTGGTAAAGGTATAATTATTGAGAATTCAAATACTACTTTTTTAAAACCGGTAGCTACGGGAAATCAAGATTTAAAAGATGGAGGTTTTGCTTTTCCTCCAACAAATCCTCCTATGTCACCAATGACATTAAATGGTATGAGAGATTTTTATAAAAATAATGAATATGTAAAAAATTTAGATGAATTGACTTTA |
| ama1.33 | ATTTGGTAAAGGTATAATTATTGAGAATTCAAAAACTACTTTTTTAACACCGGTAGCTACGGAAAATCAAGATTTAAAAGATGGAGGTTTTGCTTTTCCTCCAACAGAACCTCCTATGTCACCAATGACATTAGATGATATGAGACTTTTGTATAAAGATAATGAAGATGTAAAAAATTTAGATGAATTGACTTTA |
| ama1.34 | ATTTGGTAAAGGTATAATTATTGAGAATTCAAAAACTACTTTTTTAACACCGGTAGCTACGGAAAATCAAGATTTAAAAGATGGAGGTTTTGCTTTTCCTCCAACAGAACCTCTTATATCACCAATGACATTAGATCAAATGAGAGATTTTTATAAAAATAATGAATATGTAAAAAATTTAGATGAATTGACTTTA |
| ama1.35 | ATTTGGTAAAGGTATAATTATTGAGAATTCAAATACTACTTTTTTAACACCGGTAGCTACGGGAAATCAAGATTTAAAAGATGGAGGTTTTGCTTTTCCTCCAACAAATCCTCCTATGTCACCAATGACATTAAATGGTATGAGAGATTTATATAAAAATAATGAATATGTAAAAAATTTAGATGAATTGACTTTA |
| ama1.36 | ATTTGGTAAAGGTATAATTATTGAGAATTCAAATACTACTTTTTTAACACCGGTAGCTACGGGAAATCAAGATTTAAAAGATGGAGGTTTTGCTTTTCCTCCAACAAATCCTCCTATGTCACCAATGACATTAAATGGTATGAGAGATTTTTATAAAAATAATGAATATGTAAAAAATTTAGATGAATTGACTTTA |

**Table S5. Haplotypes detected for Heome A in 100 Cameroonian participants**

| Haplotype Name | Haplotype Sequence |
|---|---|
| heome-a.00 | TTATTAATGTCATTTTGTTTTTCTTTATTATTATTATTTTTATTTTTTCCCTGGTTATCTTCCTTATTACCTAGCTTTTTTTTGTTGTCATTATTTTCTGTCTGATTATTATTTTTGACTTTGTTCATCTTTGAATTATCCGTTCTGTTTGTATGTCTCAAGCAAGGTT |
| heome-a.01 | TTATTAATGTCATTTTGTTTTTCTTTATTATTATTATTTTTATTTTTTCCCTGGTTATCTTCCTTATTACCTAGCTTTTTTTTGTTGTCATTATTTTCTGTCTGATTATTATTTTTGACTTTGTTCATCTTTGAATTATTCGTTCTGTTTGTATGTCTCAAGCAAGGTT |
| heome-a.02 | TTATTAATGTCATTTTGTTTTTCTTTATTATTATTATTTTTATTTTTTCCCTGGTTATCTTCCTTATTACCTAGCTTTTTTTTGTTGTCATTATTTTCTGTCTGATTATTATTTTTGACTCTGTTCATCTTTGAATTATCCGTTCTGTTTGTATGTCTCAAGCAAGGTT |
| heome-a.03 | TTATTAATGTCATTTTGTTTTTCTTTATTATTATTATTTTTATTTTTTCCCTGGTTATCTTCCTTATTACCTAGCTTTTTTTTGTTGTCATTATTTTCTGTCTGATTATTATTTTTGACTTTGTTCATCCTTGAATTATCCGTTCTGTTTGTATGTCTCAAGCAAGGTT |
| heome-a.04 | TTATTAATGTCATTTTGTTTTTCTTTATTATTATTATTTTTATTTTTTCCCTGGTTATCTTTCTTATTACCTAGCTTTTTTTTGTTGTCATTATTTTCTGTCTGATTATTATTTTTGACTTTGTTCATCTTTGAATTATCCGTTCTGTTTGTATGTCTCAAGCAAGGTT |
| heome-a.05 | TTATTAATGTCATTTTGTTTTTCTTTATTATTATTATTTTTATTTTTTCCCTGGTTATCTTTCTTATTACCTAGCTTTTTTTTGTTGTCATTATTTTCTGTCTGATTATTATTTTTAACTTTGTTCATCTTTGAATTATGCGTTCTGTTTGTATGTCTCAAGCAAGGTT |
| heome-a.06 | TTATTAATGTCATTTTGTTTTTCTTTATTATTATTATTTTTATTTTTTCCCTGGTTATCTTCCTTATTACCTAGCTTTTTTTTGTTGTCATTATTTTCTGTCTGATTATTATTTTTGACTTTGTTCATCTTTGAATTATGCGTTCTGTTTGTATGTCTCAAGCAAGGTT |
| heome-a.07 | TTATTAATGTCATTTTGTTTTTCTTTATTATTATTATTTTTATTTTTTCCCTGGTTATCTTCCTTATTACCTAGCTTTTTTTTGTTGTCATTATTTTCTGTCTGATTATTATTTTTGACTTTTTTCATCTTTGAATTATCCGTTCTGTTTGTATGTCTCAAGCAAGGTT |
| heome-a.08 | TTATTAATGTCATTTTGTTTTTCTTTATTATTATTATTTTTATTTTTTCCCTGGTTATCTTTCTTATTACCTAGCTTTTTTTTGTTGTCATTATTTTCTGTCTGATTATTATTTTTGACTCTGTTCATCTTTGAATTATCCGTTCTGTTTGTATGTCTCAAGCAAGGTT |
| heome-a.09 | TTATTAATGTCATTTTGTTTTTCTTTATTATTATTATTTTTATTTTTTCCCTGGTTATCTTTCTTATTACCTAGCTTTTTTTTGTTGTCATTATTTTCTGTCTGATTATTATTTTTGACTTTGTTCATCTTTGAATTATTCGTTCTGTTTGTATGTCTCAAGCAAGGTT |
| heome-a.10 | TTATTAATGTCATTTTGTTTTTCTTTATTATTATTATTTTTATTTTTTCCCTGGTTATCTTCCTTATTACCTAGCTTTTTTTTGTTGTCATTATTTTCTGTCTGATTATTATTTTTGACTCTGTTCATCTTTGAATTATTCGTTCTGTTTGTATGTCTCAAGCAAGGTT |
| heome-a.11 | TTATTAATGTCATTTTGTTTTTCTTTATTATTATTATTTTTATTTTTTCCCTGGTTATCTTCCTTATTACCTAGCTTTTTTTTGTTGTCATTATTTTCTGTCTGATTATTATTTTTGACTTTGTTCATCTTTGAATTAACCGTTCTGTTTGTATGTCTCAAGCAAGGTT |
| heome-a.12 | TTATTAATGTCATTTTGTTTTTCTTTATTATTATTATTTTTATTTTTTCCCTGGTTATCTTCCTTATTACCTAGCTTTTTTTTGTTGTCATTATTTTCTGTCTGATTATTATTTTTGACTCTGTTCATCTTTGAATTATGCGTTCTGTTTGTATGTCTCAAGCAAGGTT |
| heome-a.13 | TTATTAATGTCATTTTGTTTTTCTTTATTATTATTATTTTTATTTTTTCCCTGGTTATCTTCCTTATTACCTAGCTTTTTTTTGTTGTCATTATTTTCTGTCTGATTATTATTTTTGACTTTGTTAATCTTTGAATTATCCGTTCTGTTTGTATGTCTCAAGCAAGGTT |
| heome-a.14 | TTATTAATGTCATTTTGTTTTTCTTTATTATTATTATTTTTATTTTTTCCCTGGTTATCTTTCTTATTACCTAGCTTTTTTTTGTTGTCATTATTTTCTGTCTGATTATTATTTTTGACTCTGTTCATCTTTGAATTATTCGTTCTGTTTGTATGTCTCAAGCAAGGTT |
| heome-a.15 | TTATTAATGTCATTTTGTTTTTCTTTATTATTATTATTTTTATTTTTTCCCTGGTTATCTTTCTTATTACCTAGCTTTTTTTTGTTGTCATTATTTTCTGTCTGATTATTATTTTTGACTTTGTTCATCCTTGAATTATCCGTTCTGTTTGTATGTCTCAAGCAAGGTT |
| heome-a.16 | TTATTAATGTCATTTTGTTTTTCTTTATTATTATTATTTTTATTTTTTCCCTGGTTATCTTCCTTATTACCTAGCTTTTTTTTGTTGTCATTATTTTCTGTCTGATTATTATTTTTGACTTTTTTCATCTTTGAATTATCCATTCTGTTTGTATGTCTCAAGCAAGGTT |
| heome-a.17 | TTATTAATGTCATTTTGTTTTTCTTTATTATTATTATTTTTATTTTTTCCCTGGTTATCTTTCTTATTACCTAGCTTTTTTTTGTTGTCATTATTTTCTGTCTGATTATTATTTTTGACTTTGTTCATCTTTGAATTAACCGTTCTGTTTGTATGTCTCAAGCAAGGTT |

| | |
|---|---|
| heome-a.18 | TTATTAATGTCATTTTGTTTTTCTTTATTATTATTATTTTTATTTTTTCCCTGGTTATCTTCCTTATTACCTAGCTTTTTTTTGTTGTCATTATTTTCTGTCTGATTATTATTTTTGACTTTGTTCATCTTTGAATTATCCGTTCTGTTTGAATGTCTCAAGCAAGGTT |

**Table S6. Haplotypes detected for Heome B in 100 Cameroonian participants**

| Haplotype Name | Haplotype Sequence |
|---|---|
| heome-b.00 | AATGTACTTATCCCCTGATATGATTTTTCCAATATCATCAAAATCAGTAATCTCTAAATCATTTATATGATTTATATGTATAAATTCTTTTATCTTGTTCAAATAATTTCTGTAATAAAATATATTCTCTTTATCATCATCAATTATTTTCATATAATCC |
| heome-b.01 | AATGTACTTATCCCCTGATATGATTTCTCCAGTATCATCAAAATCAGTAATCTCTAAATCATTTATATGATTTATATGTATAAATTCTTTTATCTTGTTCAAATAATTTCTGTAATAAAATATATTCTCTTTATCATCATCAATTATTTTCATATAATCC |
| heome-b.02 | AATGTACTTATCCCCTGATATGATTCCTCCAATATCATCAAAATCAGTAATCTCTAAATCATTTATATGATTTATATGTATAAATTCTTTTATCTTGTTCAAATAATTTCTGTAATAAAATATATTCTCTTTATCATCATCAATTATTTTCATATAATCC |
| heome-b.03 | AATGTACTTATCCCCTGATATGATTGTTCCAATATCATCAAAATCAGTAATCTCTAAATCATTTATATGATTTATATGTATAAATTCTTTTATCTTGTTCAAATAATTTCTGTAATAAAATATATTCTCTTTATCATCATCAATTATTTTCATATAATCC |
| heome-b.04 | AATGTACTTATCCCCTGATATGATTTCCCCAATATCATCAAAATCAGTAATCTCTAAATCATTTATATGATTTATATGTATAAATTCTTTTATCTTGTTCAAATAATTTCTGTAATAAAATATATTCTCTTTATCATCATCAATTATTTTCATATAATCC |
| heome-b.05 | AATGTACTTATCCCCTGATATGATTCCTCCAATATCATCAAAATTAGTAATCTCTAAATCATTTATATGATTTATATGTATAAATTCTTTTATCTTGTTCAAATAATTTCTGTAATAAAATATATTCTCTTTATCATCATCAATTATTTTCATATAATCC |
| heome-b.06 | AATGTACTTATCCCCTGATATGATTTTTCCAATATCATCAAAATTAGTAATCTCTAAATCATTTATATGATTTATATGTATAAATTCTTTTATCTTGTTCAAATAATTTCTGTAATAAAATATATTCTCTTTATCATCATCAATTATTTTCATATAATCC |
| heome-b.07 | AATGTACTTATCCCCTGATATGATTTCTCCAATATCATCAAAATCAGTAATCTCTAAATCATTTATATGATTTATATGTATAAATTCTTTTATCTTGTTCAAATAATTTCTGTAATAAAATATATTCTCTTTATCATCATCAATTATTTTCATATAATCC |
| heome-b.08 | AATGTACTTATCCCCTGATATGATTTCTCCATTATCATCAAAATCAGTAATCTCTAAATCATTTATATGATTTATATGTATAAATTCTTTTATCTTGTTCAAATAATTTCTGTAATAAAATATATTCTCTTTATCATCATCAATTATTTTCATATAATCC |
| heome-b.09 | AATGTACTTATCCCCTGATATGAGTTCTCCAATATCATCAAAATCAGTAATCTCTAAATCATTTATATGATTTATATGTATAAATTCTTTTATCTTGTTCAAATAATTTCTGTAATAAAATATATTCTCTTTATCATCATCAATTATTTTCATATAATCC |

**Table S7. Haplotypes detected for Heome C in 100 Cameroonian participants**

| Haplotype Name | Haplotype Sequence |
|---|---|
| heome-c.0 | TACCCACGGGACATTTTAATTTATACCTATTGGAACATTTTTTATTAGGTCTTTCTTTATGATTCTTCTTTAAAATAGGTTCATCATTATTTGAGTAAACTAACAGATCATTATCGTTCT<br>CAAAATTATTATAACCATCAATATTTTTATTTCCATATACATCTATAAT |
| heome-c.1 | TACCCACGGGACATTTTAATTTATACCTATTGGAACATTTTTTATTAGGTCTTTCTTTATGATTCTTCTTTAAAATAGGTTCATCATTATTTGAGTAAACTAACAGATTATTATCGTTCT<br>CAAAATTATTATAACCATCAATATTTTTATTTCCATATACATCTATAAT |
| heome-c.2 | TACCCACGGGACATTTTAATTTATACCTATTGGAACATTTTTTATTAGGTCTTTCTTTATGATTCTTCTTTAAAATAGGTTCATCATTATTTGAGTAAACTAACAGATCATTATCGTTCT<br>CAAATTTATTATAACCATCAATATTTTTATTTCCATATACATCTATAAT |
| heome-c.3 | TACCCACGGGACATTTTAATTTATACCTATTGGAACATTTTTTATTAGGTCTTTCTTTATGATTCTTCTTTAAAATAGGTTCATCATTATTTGAGTAAACTAACAGATCATTATGGTTC<br>TCAAAATTATTATAACCATCAATATTTTTATTTCCATATACATCTATAAT |
| heome-c.4 | TACCCACGGGACATTTTAATTTATACCTATTGGAACATTTTTTATTAGGTCTTTCTTTATGATTCTTCTTTAAAATAGGTTCATCATTATTTGAGTAAACTAACAGATCATTATCCTTCT<br>CAAAATTATTATAACCATCAATATTTTTATTTCCATATACATCTATAAT |

**Table S8. Haplotypes detected for Heome D in 100 Cameroonian participants**

| Haplotype Name | Haplotype Sequence |
|---|---|
| heome-d.00 | CATGATTCCTCAAAAGTTTAACATTCCACATAGTATCTGTTAAATTGATAGTTTGCAAACAATTATACGTTTGGTTATTTTTATTTTTAAAAAAGGATATATGATCATCTAAAGATTTATTTTTGAGAAAGATTTCCATATATTCATATATATCTTGAATACACCAG |
| heome-d.01 | CATGATTCCTCAAAAGTTTAACATTCCACATAGTATCTGTTAAATTGATAGTTTGCAAACAATTATACGTTTGGTTATTTTTATTTTTAAAAAAGGATATATGATCATCTAAAGATTTATTTTTGAGAAAGATTTCCATATATTCATATATATCATGAATACACCAG |
| heome-d.02 | CATGATTCCTCAAAAGTTTAACATTCCACATAGTATCTGTTAAATTGATAGTTTGCAAACAATTATACGTTTGGTTATTTTTATTTTTAAAAAAGGATATATGATCATCTAAAGATTTATTTTTGAGAAAGATTTCCATATATTCATATATATCTTGGATACACCAG |
| heome-d.03 | CATGATTCCTCAAAAGTTTAACATTCCACATAGTATCTGTTAAATTGATAGTTTGCAAACAATTATACGTTTGGTTATTTTTATTTTTAAAAAAGGATATATGATCATCTAAACATTTATTTTTGAGAAAGATTTCCATATATTCATATATATCTTGAATACACCAG |
| heome-d.04 | CATGATTCCTCAAAAGTTTAACATTCCACATAGTATCTGTTAAATTGATAGTTTGCAAACAATTATACGTTTGGTTATTTTTATTTTTAAAAAAGGATATATGATCATCTAAACATTTATTTTTGAGAAAGATTTCCATATATTCATATATATCTTGGATACACCAG |
| heome-d.05 | CATGATTCCTCAAAAGTTTAACATTCCACATAGTATCTGTTAAATTGATAGTTTGCAAACAATTATACGTTTGGTTATTTTTATTTTTAAAAAAGGATATATGATCATCTAACGATTTATTTTTGAGAAAGATTTCCATATATTCATATATATCTTGAATACACCAG |
| heome-d.06 | CATGATTCCTCAAAAGTTTAACATTCCACATAGTATCTGTTAAATTGATAGTTTGCAAACAATTATACGTTTGGTTATTTTTATTTTTAAAAAAGGATATATGATCATCTAAAGATTTATTTTTGAGAAAGATTTCCATATATTCATATACATCTTGAATACACCAG |
| heome-d.07 | CATGATTCCTCAAAAGTTTAACATTCCACATAGTATCTGTTAAATTGATAGTTTGCAAACAATTATACGTTTGGTTATTTTTATTTTTAAAAAAGGATATATGATCATCTAAAGATTTATTTTTGAGAAAGATTTCCATATATTCATATATATCTTGTATACACCAG |
| heome-d.08 | CATGATTCCTCAAAAGTTTAACATTCCACATAGTATCTGTTAAATTGATAGTTTGCAAACAATTATACGTTTGGTTATTTTTATTTTTAAAAAAGGATATATGATCATCTAAACATTTATTTTTGAGAAAGATTTCCATATATTCATATACATCTTGAATACACCAG |
| heome-d.09 | CATGATTCCTCAAAAGTTTAACATTCCACATAGTATCTGTTAAATTGATAGTTTGCAAACAATTATACGTTTGGTTATTTTTATTTTTAAAAAAGGATATATGATCATCTAAAGATTTATTTTTGAGAAAGATTTCCATATATTCATATATATCGTGAATACACCAG |
| heome-d.10 | CATGATTCCTCAAAAGTTTAACATTCCACATAGTATCTGTTAAATTGATAGTTTGCAAACAATTATACGTTTGGTTATTTTTATTTTTAAAAAAGGATATATGATCATCTAAAAATTTATTTTTGAGAAAGATTTCCATATATTCATATATATCTTGAATACACCAG |
| heome-d.11 | CATGATTCCTCAAAAGTTTAACATTCCACATAGTATCTGTTAAATTGATAGTTTGCAAACAATTATACCTTTGGTTATTTTTATTTTTAAAAAAGGATATATGATCATCTAAAGATTTATTTTTGAGAAAGATTTCCATATATTCATATATATCTTGAATACACCAG |
| heome-d.12 | CATGATTCCTCAAAAGTTTAACATTCCACATAGTATCTGTTAAATTGATAGTTTGCAAACAATTATACGTTTGGTTATTTTTATTTTTGAAAAAGGATATATGATCATCTAAAGATTTATTTTTGAGAAAGATTTCCATATATTCATATATATCTTGAATACACCAG |
| heome-d.13 | CATGATTCCTCAAAAGTTTAACATTCCACATAGTATCTGTTAAATTGATAGTTTGCAAACAATTATACGTTTGGTTATTTTTATTTTTAAAAAAGGATATATCATCATCTAAAGATTTATTTTTGAGAAAGATTTCCATATATTCATATATATCTTGAATACACCAG |
| heome-d.14 | CATGATTCCTCAAAAGTTTAACATTCCACATAGTATCTGTTAAATTGATAGTTTGCAAACAATTATACGTTTGGTTATTTTTATTTTTAAAAAAGGATATATGATCATCTAAAGATTTATTTTTGAAAAAGATTTCCATATATTCATATACATCTTGAATACACCAG |
| heome-d.15 | CATGATTCCTCAAAAGTTTAACATTCCACATAGTATCTGTTAAATTGATAGTTTGCAAACAATTATACCTTTGGTTATTTTTATTTTTAAAAAAGGATATATGATCATCTAAAGATTTATTTTTGAAAAAGATTTCCATATATTCATATATATCTTGAATACACCAG |
| heome-d.16 | CATGATTCCTCAAAAGTTTAACATTCCACATAGTATCTGTTAAATTGATAGTTTGCAAACAATTATACGTTTCGTTATTTTTATTTTTAAAAAAGGATATATGATCATCTAAAGATTTATTTTTGAGAAAGATTTCCATATATTCATATATATCTTGAATACCCCAG |
| heome-d.17 | CATGATTCCTCAAAAGTTTAACATTCCACATAGTATCTGTTAAATTGATAGTTTGCAAACAATTATACGTTTGGTTATTTTTATTTTTGAAAAAGGATATATGATCATCTAAAGATTTATTTTTGAGAAAGATTTCCATATATTCATATATATCATGAATACACCAG |

| | |
|---|---|
| heome-d.18 | CATGATTCCTCAAAAGTTTAACATTCCACATAGTATCTGTTAAATTGATAGTTTGCAAACAATTATACGTTTGGTTATTTTTATTTTTAAAAAAGGATATATGATCATCTAAAGATTTATTTTTAAGAAAGATTTCCATATATTCATATATATCTTGAATACACCAG |
| heome-d.19 | CATGATTCCTCAAAAGTTTAACATTCCACATAGTATCTGTTAAATTGATAGTTTGCAAACAATTATACGTTTGGTTATTTTTATTTTTAAAAAAGGATATATGATCATCTAAAGATTTATTTTTAAGAAAGATTTCCATATATTCATATATATCATGAATACACCAG |
| heome-d.20 | CATGATTCCTCAAAAGTTTAACATTCCACATAGTATCTGTTAAATTGATAGTTTGCAAACAATTATACGTTTGGTTATTTTTATTTTTGAAAAAGGATATATGATCATCTAAAGATTTATTTTTGAGAAAGATTTCCATATATTCATATATATCTTGGATACACCAG |
| heome-d.21 | CATGATTCCTCAAAAGTTTAACATTCCACATAGTATCTGTTAAATTGATAGTTTGCAAACAATTATACGTTTGGTTATTTTTATTTTTAAAAAAGGATATATGATCATCTAAACATTTATTTTTAAGAAAGATTTCCATATATTCATATATATCTTGAATACACCAG |
| heome-d.22 | CATGATTCCTCAAAAGTTTAACATTCCACATAGTATCTGTTAAATTGATAGTTTGCAAACAATTATACGTTTGGTTATTTTTATTTTTAAAAAAGGATATATGATCATCTAAAGATTTATTTTTCAGAAAGATTTCCATATATTCATATATATCATGAATACACCAG |
| heome-d.23 | CATGATTCCTCAAAAGTTTAACATTCCACATAGTATCTGTTAAATTGATAGTTTGCAAACAATTATACGTTTCGTTATTTTTATTTTTAAAAAAGGATATATGATCATCTAAAGATTTATTTTTGAGAAAGATTTCCATATATTCATATATATCTTGAATACACCAG |
| heome-d.24 | CATGATTCCTCAAAAGTTTAACATTCCACATAGTATCTGTTAAATTGATAGTTTGTAAACAATTATACGTTTGGTTATTTTTATTTTTAAAAAAGGATATATGATCATCTAAAGATTTATTTTTGAGAAAGATTTCCATATATTCATATATATCTTGGATACACCAG |
| heome-d.25 | CATGATTCCTCAAAAGTTTAACATTCCACATAGTATCTGTTAAATTGATAGTTTGCAAACAATTATACGTTTCGTTATTTTTATTTTTAAAAAAGGATATATGATCATCTAAAGATTTATTTTTGAGAAAGATTTCCATATATTCATATATATCTTGGATACACCAG |
| heome-d.26 | CATGATTCCTCAAAAGTTTAACATTCCACATAGTATCTGTTAAATTGATAGTTTGCAAACAATTATACCTTTGGTTATTTTTATTTTTAAAAAAGGATATATGATCATCTAAAGATTTATTTTTGAGAAAGATTTCCATATATTCATATATATCATGAATACACCAG |
| heome-d.27 | CATGATTCCTCAAAAGTTTAACATTCCACATAGTATCTGTTAAATTGATAGTTTGCAAACAATTATACGCTTGGTTATTTTTATTTTTAAAAAAGGATATATGATCATCTAAAGATTTATTTTTGAGAAAGATTTCCATATATTCATATATATCTTGGATACACCAG |
| heome-d.28 | CATGATTCCTCAAAAGTTTAACATTCCACATAGTATCTGTTAAATTGATAGTTTGCAAACAATTATACGTTTGGTTATTTTTATTTTTGAAAAAGGATATATGATCATCTAAAGATTTATTTTTGAGAAAGATTTCCATATATTCATATATATCGTGAATACACCAG |

**Table S9. Haplotypes detected for Heome E in 100 Cameroonian participants**

| Haplotype Name | Haplotype Sequence |
|---|---|
| heome-e.0 | GTATCTTTTCCATATATAATAACGCATGAATTCTTTAAATATATCTTTTACTTTATCTGTAAGTTTCATGGAATCTATTATTTCATGTGCATATTTTATTTTTGCCTCGTTTATATATTTACCTGCTTCTAATTTACATAAACATTTTTCTATTAAGTTTTGA |
| heome-e.1 | GTATCTTTTCCATATATAATAACGCATGAATTCTTTAAATATATCTTTTACTTTATCTGTAAGTTTCATGGAATCTATTATTTCATGTGCATATTTTATTTTTGCCTCGTTTATATATTTACCTGCTTTTAATTTACATAAACATTTTTCTATTAAGTTTTGA |
| heome-e.2 | GTATCTTTTCCATATATAATAACGCATGAATTCTTTAAATATATCTTTTACTTTATCTGTAAGTTTCATGGAATCTATTATTTCATCTGCATATTTTATTTTTGCCTCGTTTATATATTTACCTGCTTCTAATTTACATAAACATTTTTCTATTAAGTTTTGA |
| heome-e.3 | GTATCTTTTCCATATATAATAACGCATGAATTCTTTAAATATATCTTTTACTTTATCTGTAAGTTTCATGGAATCTATTATTTCTTCTGCATATTTTATTTTTGCCTCGTTTATATATTTACCTGCTTCTAATTTACATAAACATTTTTCTATTAAGTTTTGA |
| heome-e.4 | GTATCTTTTCCATATATAATAACGCATGAATTCTTTAAATATATCTTTTACTTTATCTGTAAGTTTCATGGAATCTATTATTTCATCTGCATATTTTATTTTTGCCTCGTTTATATATTTACCTGCTTTTAATTTACATAAACATTTTTCTATTAAGTTTTGA |
| heome-e.5 | GTATCTTTTCCATATATAATAACGCATGAATTCTTTAAATTTATCTTTTACTTTATCTGTAAGTTTCATGGAATCTATTATTTCATCTGCATATTTTATTTTTGCCTCGTTTATATATTTACCTGCTTCTAATTTACATAAACATTTTTCTATTAAGTTTTGA |
| heome-e.6 | GTATCTTTTCCATATATAATAACGCATGAATTCTTTAAATATATCTTTTACTTTATCTGTAAGTTTCATGGAATCTATTATTTCTTCTGCATATTTTATTTTTGCCTCGTTTATATATTTACCTGCTTTTAATTTACATAAACATTTTTCTATTAAGTTTTGA |
| heome-e.7 | GTATCTTTTCCATATATAATAACGCATGAATTCTTTAAATATATCTTTTACTTTATCTGTAAGTTTCATGGAATCTATTATTTCATGTGCATATTTTATTTTTGCCTCGTTTATATATTCACCTGCTTTTAATTTACATAAACATTTTTCTATTAAGTTTTGA |

### Table S10. Haplotypes detected for Heome F in 100 Cameroonian participants

| Haplotype Name | Haplotype Sequence |
|---|---|
| heome-f.00 | TTTGGGTAGATATCGTTATAAGGGCTATAATGTATTGACTTCTCCGGGTTTGTTACATTATCATTACAACCAATATTATTAAGGTTAATATTATTTTTATCATTATTATTGTTGTTTTCAT<br>TTGTGTGGTTATATAATAACAATGATGTGTCGTCTATTGGATCATTT |
| heome-f.01 | TTTGGGTAGATATCGTTATAAGGGCTATAATGTATTGACTTCTCCGGGTTGGTTACATTATCATTACAACCAATATTATTAAGGTTAATATTATTTTTATCATTATTATTGTTGTTTTCAT<br>TTGTGTGGTTATATAATAACAATGATGTGTCGTCTATTGGATCATTT |
| heome-f.02 | TTTGGGTAGATATCGTTATAAGGGCTATAATGTATTGACTTCTCCGGGTTGGTTACATTATCATTACAACCAATATTATTAAGGTTCATATTATTTTTATCATTATTATTGTTGTTTTCA<br>TTTGTGTGGTTATATAATAACAATGATGTGTCGTCTATTGGATCATTT |
| heome-f.03 | TTTGGGTAGATATCGTTATAAGGGCTATAATGTATTGACTTCTCCGGGTTTGTTACATTATCATTACAACCAATATTATTAAGGTTCATATTATTTTTATCATTATTATTGTTGTTTTCAT<br>TTGTGTGGTTATATAATAACAATGATGTGTCGTCTATTGGATCATTT |
| heome-f.04 | TTTGGGTAGATATCGTTATAAGGGCTATAATGTATTGACTTCTCCGGGTTTGTTACATTATCATTACAACCAATATTATTAAGGTTAATACTATTTTTATCATTATTATTGTTGTTTTCAT<br>TTGTGTGGTTATATAATAACAATGATGTGTCGTCTATTGGATCATTT |
| heome-f.05 | TTTGGGTAGATATCGTTATAAGGGCTATAATGTATTGACTTCTCCGGGTTGGTTACATTATCATTACAACCAATATTATTAAGGTTAATACTATTTTTATCATTATTATTGTTGTTTTCA<br>TTTGTGTGGTTATATAATAACAATGATGTGTCGTCTATTGGATCATTT |
| heome-f.06 | TTTGGGTAGATATCGTTATAAGGGCTATAATGTATTGACTTCTCCGGGTTTGTTACATTATCATTACAACCAATATTATTAAGGTTCATATTATTTTTATCATTATTATTCTTGTTTTCAT<br>TTGTGTGGTTATATAATAACAATGATGTGTCGTCTATTGGATCATTT |
| heome-f.07 | TTTGGGTAGATATCGTTATAAGGGCTATAATGTATTGACTTCTCCGGGTTTGTTACATTATCATTACAACCAATATTATTAAGGTTAATATTATTTTTACCATTATTATTGTTGTTTTCAT<br>TTGTGTGGTTATATAATAACAATGATGTGTCGTCTATTGGATCATTT |
| heome-f.08 | TTTGGGTAGATATCGTTATAAGGGCTATAATGTATTGACTTCTCCGGGTTTGTTACATTATCATTACAACCAATATTATTAAGGTTAATATTATTTTTATCATTATTATTCTTGTTTTCAT<br>TTGTGTGGTTATATAATAACAATGATGTGTCGTCTATTGGATCATTT |
| heome-f.09 | TTTGGGTAGATATCGTTATAAGGGCTATAATGTATTGACTTCTCCGGGTTTGTTACATTATCATTACAACCAATATTATTAAGGTTAATATTATTTTTATCATTATTATTGTTGTTTTCAT<br>TTGTGTGGTTATATAATAACAATGATGTGTCGTCTATTAGATCATTT |
| heome-f.10 | TTTGGGTAGATATCGTTATAAGGGATATAATGTATTGACTTCTCCGGGTTGGTTACATTATCATTACAACCAATATTATTAAGGTTCATATTATTTTTATCATTATTATTGTTGTTTTCAT<br>TTGTGTGGTTATATAATAACAATGATGTGTCGTCTATTGGATCATTT |
| heome-f.11 | TTTGGGTAGATATCGTTATAAGGGCTATAATGTATTGACTTCTCCGGGTTGGTTACATTATCATTACAACCAATATTATTAAGGTTAATATTATTTTTACCATTATTATTGTTGTTTTCA<br>TTTGTGTGGTTATATAATAACAATGATGTGTCGTCTATTGGATCATTT |
| heome-f.12 | TTTGGGTAGATATCGTTATAAGGGCTATAATGTATTGACTTCTCCGGGTTGGTTACATTATCATTACAACCAATATTATTAAGGTTAATATTATTTTTATCATTATTATTCTTGTTTTCAT<br>TTGTGTGGTTATATAATAACAATGATGTGTCGTCTATTGGATCATTT |

**Table S11. Haplotypes detected for Heome G in 100 Cameroonian participants**

| Haplotype Name | Haplotype Sequence |
|---|---|
| heome-g.0 | AATGATGTAGATGCTAACACAACATCCTGGTCAGTATCTTGTGATAAATAATTTAATTTGTCCGCTAAAAAATAAAGTATAATTTCACAGCTTTTTTTATTTGTATCTGTGTTTTTCATGTTATCATAAAAAGACAAAGATGAAGTAGCACATGAGTTAATTAT |
| heome-g.1 | AATGATGTAGATGCTAATAGCACATCCTGGTCAGTATCTTGTGATAAATAATTTAATTTATCCGCTAAAAAATAAAGTATAATTTCACAGCTTTTTTTATTTGTATCTGTGTTTTTCATGTTATCATAAAAAGACAAAGATGAAGTAGCACATGAGTTAATTAT |
| heome-g.2 | AATGATGTAGATGCTAACACAACATCCTGGTCAGTATCTTGTGATAAATAATTTAATTTATCCGCTAAAAAATAAAGTATAATTTCACAGCTTTTTTTATTTGTATCTGTGTTTTTCATGTTATCATAAAAAGACAAAGATGAAGTAGCACATGAGTTAATTAT |
| heome-g.3 | AATGATGTAGATGCTAATAGCACATCCTGGTCAGTATCTTGTGATAAATAATTTAATTTGTCCGCTAAAAAATAAAGTATAATTTCACAGCTTTTTTTATTTGTATCTGTGTTTTTCATGTTATCATAAAAAGACAAAGATGAAGTAGCACATGAGTTAATTAT |
| heome-g.4 | AATGATGTAGATGCTAACACAACATCCTGGTCAGTATCTTGTGATAAATAATTTAATTTGTCCGCTAAAAAATAAAGTATAATTTCACAGCTTTTTTTATTTGTATCTGTGTTTTTCATGTTATCGTAAAAAGACAAAGATGAAGTAGCACATGAGTTAATTAT |
| heome-g.5 | AATGATGTAGATGCTAACACAACATCCTGGTCAGTATCTTGTGATAAATAATTTAATTTATCCGATAAAAAATAAAGTATAATTTCACAGCTTTTTTTATTTGTATCTGTGTTTTTCATGTTATCATAAAAAGACAAAGATGAAGTAGCACATGAGTTAATTAT |

**Table S12. Haplotypes detected for Heome H in 100 Cameroonian participants**

| Haplotype Name | Haplotype Sequence |
|---|---|
| heome-h.0 | AAGAGTGTTAATAATACTACAGCTAGTAATATTTCCAAATCCAAAAATGAACTTGAAAAAAAGTCATCAGTTAATAATAATGCTTATATTAAAATTATCGAAAATTATGATTTATTATGGTTAAAATTAATAGAAATTGAATTATGTTGTAATAATAAAAATTTAAATCCTATAA |
| heome-h.1 | AAGAGTGTTAATAATACTACAGCTAGTCATATTTCCAAATCCAAAAATGAACTTGAAAAAAAGTCATCAGCTAATAATAATGCTTATATTAAAATAATCGAAAATTATGATTTATTATGGTTAAAATTAATAGAAATTGAATTATGTTGTAATAATAAAAATTTAAATCCTATAA |
| heome-h.2 | AAGAGTGTTAATAATACTACAGCTAGTCATATTTCCAAATCCAAAAATGAACTTGAAAAAAAGTCATCAGCTAATAATAATGCTTATATTAAAATTATCGAAAATTATGATTTATTATGGTTAAAATTAATAGAAATTGAATTATGTTGTAATAATAAAAATTTAAATCCTATAA |
| heome-h.3 | AAGAGTGTTAATAATACTACAGCTAGTCATATTTCCAAATCCAAAAATGAACTTGAAAAAAAGTCATCAGTTAATAATAATGCTTATATTAAAATTATCGAAAATTATGATTTATTATGGTTAAAATTAATAGAAATTGAATTATGTTGTAATAATAAAAATTTAAATCCTATAA |
| heome-h.4 | AAGAGTGTTAATAATACTACAGCTAGTAATATTTCCAAATCCAAAAATGAACTTGAAAAAAAGTCATCAGCTAATAATAATGCTTATATTAAAATAATCGAAAATTATGATTTATTATGGTTAAAATTAATAGAAATTGAATTATGTTGTAATAATAAAAATTTAAATCCTATAA |

**Table S13. ITrue Index ordering**

| Primer Name | Primer Sequence |
|---|---|
| iTru5_01_A | AATGATACGGCGACCACCGAGATCTACACACCGACAAACACTCTTTCCCTA*C |
| iTru5_01_B | AATGATACGGCGACCACCGAGATCTACACAGTGGCAAACACTCTTTCCCTA*C |
| iTru5_01_C | AATGATACGGCGACCACCGAGATCTACACCACAGACTACACTCTTTCCCTA*C |
| iTru5_01_D | AATGATACGGCGACCACCGAGATCTACACCGACACTTACACTCTTTCCCTA*C |
| iTru5_01_E | AATGATACGGCGACCACCGAGATCTACACGACTTGTGACACTCTTTCCCTA*C |
| iTru5_01_F | AATGATACGGCGACCACCGAGATCTACACGTGAGACTACACTCTTTCCCTA*C |
| iTru5_01_G | AATGATACGGCGACCACCGAGATCTACACGTTCCATGACACTCTTTCCCTA*C |
| iTru5_01_H | AATGATACGGCGACCACCGAGATCTACACTAGCTGAGACACTCTTTCCCTA*C |
| iTru5_02_A | AATGATACGGCGACCACCGAGATCTACACCTTCGCAAACACTCTTTCCCTA*C |
| iTru5_02_B | AATGATACGGCGACCACCGAGATCTACACGTGGTATGACACTCTTTCCCTA*C |
| iTru5_02_C | AATGATACGGCGACCACCGAGATCTACACCACTGTAGACACTCTTTCCCTA*C |
| iTru5_02_D | AATGATACGGCGACCACCGAGATCTACACAGACGCTAACACTCTTTCCCTA*C |
| iTru5_02_E | AATGATACGGCGACCACCGAGATCTACACCAACTCCAACACTCTTTCCCTA*C |
| iTru5_02_F | AATGATACGGCGACCACCGAGATCTACACAACACGCTACACTCTTTCCCTA*C |
| iTru5_02_G | AATGATACGGCGACCACCGAGATCTACACTGGATGGTACACTCTTTCCCTA*C |
| iTru5_02_H | AATGATACGGCGACCACCGAGATCTACACTTCGAAGCACACTCTTTCCCTA*C |
| iTru5_03_A | AATGATACGGCGACCACCGAGATCTACACAACACCACACACTCTTTCCCTA*C |
| iTru5_03_B | AATGATACGGCGACCACCGAGATCTACACTGAGCTGTACACTCTTTCCCTA*C |
| iTru5_03_C | AATGATACGGCGACCACCGAGATCTACACCACAGGAAACACTCTTTCCCTA*C |
| iTru5_03_D | AATGATACGGCGACCACCGAGATCTACACTGACAACCACACTCTTTCCCTA*C |
| iTru5_03_E | AATGATACGGCGACCACCGAGATCTACACTGTTCCGTACACTCTTTCCCTA*C |
| iTru5_03_F | AATGATACGGCGACCACCGAGATCTACACCCTAGAGAACACTCTTTCCCTA*C |
| iTru5_03_G | AATGATACGGCGACCACCGAGATCTACACGCATAACGACACTCTTTCCCTA*C |
| iTru5_03_H | AATGATACGGCGACCACCGAGATCTACACCAGTGCTTACACTCTTTCCCTA*C |
| iTru5_04_A | AATGATACGGCGACCACCGAGATCTACACCGTATCTCACACTCTTTCCCTA*C |
| iTru5_04_B | AATGATACGGCGACCACCGAGATCTACACCGTCAAGAACACTCTTTCCCTA*C |
| iTru5_04_C | AATGATACGGCGACCACCGAGATCTACACCCATGAACACACTCTTTCCCTA*C |
| iTru5_04_D | AATGATACGGCGACCACCGAGATCTACACGGTACTTCACACTCTTTCCCTA*C |
| iTru5_04_E | AATGATACGGCGACCACCGAGATCTACACACCGCTATACACTCTTTCCCTA*C |
| iTru5_04_F | AATGATACGGCGACCACCGAGATCTACACTTCCAGGTACACTCTTTCCCTA*C |
| iTru5_04_G | AATGATACGGCGACCACCGAGATCTACACTCGAACCTACACTCTTTCCCTA*C |
| iTru5_04_H | AATGATACGGCGACCACCGAGATCTACACTAGTGCCAACACTCTTTCCCTA*C |
| iTru5_05_A | AATGATACGGCGACCACCGAGATCTACACGGTACGAAACACTCTTTCCCTA*C |
| iTru5_05_B | AATGATACGGCGACCACCGAGATCTACACAAGCATCGACACTCTTTCCCTA*C |
| iTru5_05_C | AATGATACGGCGACCACCGAGATCTACACGCCAATACACACTCTTTCCCTA*C |

| | |
|---|---|
| iTru5_05_D | AATGATACGGCGACCACCGAGATCTACACCTGTATGCACACTCTTTCCCTA*C |
| iTru5_05_E | AATGATACGGCGACCACCGAGATCTACACCTTAGGACACACTCTTTCCCTA*C |
| iTru5_05_F | AATGATACGGCGACCACCGAGATCTACACTCAGCCTTACACTCTTTCCCTA*C |
| iTru5_05_G | AATGATACGGCGACCACCGAGATCTACACACATGCCAACACTCTTTCCCTA*C |
| iTru5_05_H | AATGATACGGCGACCACCGAGATCTACACGATGGAGTACACTCTTTCCCTA*C |
| iTru5_06_A | AATGATACGGCGACCACCGAGATCTACACCGATCGATACACTCTTTCCCTA*C |
| iTru5_06_B | AATGATACGGCGACCACCGAGATCTACACTACTCCAGACACTCTTTCCCTA*C |
| iTru5_06_C | AATGATACGGCGACCACCGAGATCTACACAGCTACCAACACTCTTTCCCTA*C |
| iTru5_06_D | AATGATACGGCGACCACCGAGATCTACACTCGACAAGACACTCTTTCCCTA*C |
| iTru5_06_E | AATGATACGGCGACCACCGAGATCTACACTATGACCGACACTCTTTCCCTA*C |
| iTru5_06_F | AATGATACGGCGACCACCGAGATCTACACAGCCAACTACACTCTTTCCCTA*C |
| iTru5_06_G | AATGATACGGCGACCACCGAGATCTACACGATCTTGCACACTCTTTCCCTA*C |
| iTru5_06_H | AATGATACGGCGACCACCGAGATCTACACCCTCGTTAACACTCTTTCCCTA*C |
| iTru7_101_01 | CAAGCAGAAGACGGCATACGAGATGGTAACGTGTGACTGGAGTTCA*G |
| iTru7_101_02 | CAAGCAGAAGACGGCATACGAGATCAACACAGGTGACTGGAGTTCA*G |
| iTru7_101_03 | CAAGCAGAAGACGGCATACGAGATACACCTCAGTGACTGGAGTTCA*G |
| iTru7_101_04 | CAAGCAGAAGACGGCATACGAGATCATGGATCGTGACTGGAGTTCA*G |
| iTru7_101_05 | CAAGCAGAAGACGGCATACGAGATTGATAGGCGTGACTGGAGTTCA*G |
| iTru7_101_06 | CAAGCAGAAGACGGCATACGAGATCGGTTGTTGTGACTGGAGTTCA*G |
| iTru7_101_07 | CAAGCAGAAGACGGCATACGAGATCAACGAGTGTGACTGGAGTTCA*G |
| iTru7_101_08 | CAAGCAGAAGACGGCATACGAGATACCATAGGGTGACTGGAGTTCA*G |
| iTru7_101_09 | CAAGCAGAAGACGGCATACGAGATGGTGTACAGTGACTGGAGTTCA*G |
| iTru7_101_10 | CAAGCAGAAGACGGCATACGAGATCAGCATACGTGACTGGAGTTCA*G |
| iTru7_101_11 | CAAGCAGAAGACGGCATACGAGATGGACATCAGTGACTGGAGTTCA*G |
| iTru7_101_12 | CAAGCAGAAGACGGCATACGAGATAGAAGGACGTGACTGGAGTTCA*G |
| iTru7_102_01 | CAAGCAGAAGACGGCATACGAGATCGCCTTATGTGACTGGAGTTCA*G |
| iTru7_102_02 | CAAGCAGAAGACGGCATACGAGATCAGGTAAGGTGACTGGAGTTCA*G |
| iTru7_102_03 | CAAGCAGAAGACGGCATACGAGATTTGCAACGGTGACTGGAGTTCA*G |
| iTru7_102_04 | CAAGCAGAAGACGGCATACGAGATGCTGAATCGTGACTGGAGTTCA*G |
| iTru7_102_05 | CAAGCAGAAGACGGCATACGAGATGAACGTGAGTGACTGGAGTTCA*G |
| iTru7_102_06 | CAAGCAGAAGACGGCATACGAGATAACGCACAGTGACTGGAGTTCA*G |
| iTru7_102_07 | CAAGCAGAAGACGGCATACGAGATCGCAACTAGTGACTGGAGTTCA*G |
| iTru7_102_08 | CAAGCAGAAGACGGCATACGAGATTGGCTCTTGTGACTGGAGTTCA*G |
| iTru7_102_09 | CAAGCAGAAGACGGCATACGAGATTGAGCTGTGTGACTGGAGTTCA*G |
| iTru7_102_10 | CAAGCAGAAGACGGCATACGAGATGCCTTAACGTGACTGGAGTTCA*G |
| iTru7_102_11 | CAAGCAGAAGACGGCATACGAGATTGTGGCTTGTGACTGGAGTTCA*G |
| iTru7_102_12 | CAAGCAGAAGACGGCATACGAGATAACCGTGTGTGACTGGAGTTCA*G |

| | |
|---|---|
| iTru7_103_01 | CAAGCAGAAGACGGCATACGAGATAATCGCTGGTGACTGGAGTTCA*G |
| iTru7_103_02 | CAAGCAGAAGACGGCATACGAGATGGTCACTAGTGACTGGAGTTCA*G |
| iTru7_103_03 | CAAGCAGAAGACGGCATACGAGATTAGTCTCGGTGACTGGAGTTCA*G |
| iTru7_103_04 | CAAGCAGAAGACGGCATACGAGATACCATGTCGTGACTGGAGTTCA*G |
| iTru7_103_05 | CAAGCAGAAGACGGCATACGAGATAGACATGCGTGACTGGAGTTCA*G |
| iTru7_103_06 | CAAGCAGAAGACGGCATACGAGATGATGGAGTGTGACTGGAGTTCA*G |
| iTru7_103_07 | CAAGCAGAAGACGGCATACGAGATCAGTCACAGTGACTGGAGTTCA*G |
| iTru7_103_08 | CAAGCAGAAGACGGCATACGAGATGTTCTTCGGTGACTGGAGTTCA*G |
| iTru7_103_09 | CAAGCAGAAGACGGCATACGAGATAAGACACCGTGACTGGAGTTCA*G |
| iTru7_103_10 | CAAGCAGAAGACGGCATACGAGATGCCTTCTTGTGACTGGAGTTCA*G |
| iTru7_103_11 | CAAGCAGAAGACGGCATACGAGATTCGAACCTGTGACTGGAGTTCA*G |
| iTru7_103_12 | CAAGCAGAAGACGGCATACGAGATGGAACATGGTGACTGGAGTTCA*G |
| iTru7_104_01 | CAAGCAGAAGACGGCATACGAGATTATGGCACGTGACTGGAGTTCA*G |
| iTru7_104_02 | CAAGCAGAAGACGGCATACGAGATCTACAAGGGTGACTGGAGTTCA*G |
| iTru7_104_03 | CAAGCAGAAGACGGCATACGAGATAATCCAGCGTGACTGGAGTTCA*G |
| iTru7_104_04 | CAAGCAGAAGACGGCATACGAGATCCTCGTTAGTGACTGGAGTTCA*G |
| iTru7_104_05 | CAAGCAGAAGACGGCATACGAGATGCAACCATGTGACTGGAGTTCA*G |
| iTru7_104_06 | CAAGCAGAAGACGGCATACGAGATGGTATAGGGTGACTGGAGTTCA*G |
| iTru7_104_07 | CAAGCAGAAGACGGCATACGAGATCGACCTAAGTGACTGGAGTTCA*G |
| iTru7_104_08 | CAAGCAGAAGACGGCATACGAGATGATCTTGCGTGACTGGAGTTCA*G |
| iTru7_104_09 | CAAGCAGAAGACGGCATACGAGATAAGGCTCTGTGACTGGAGTTCA*G |
| iTru7_104_10 | CAAGCAGAAGACGGCATACGAGATTCCATTGCGTGACTGGAGTTCA*G |
| iTru7_104_11 | CAAGCAGAAGACGGCATACGAGATTACTCCAGGTGACTGGAGTTCA*G |
| iTru7_104_12 | CAAGCAGAAGACGGCATACGAGATCGATGTTCGTGACTGGAGTTCA*G |

### Protocol 1. *Pf*-SMARRT Amplification

***Plasmodium falciparum Simultaneous Multiplex Antimalarial Resistance and Relatedness Testing v1.9***

**Infectious Disease Epidemiology and Ecology Lab University of North Carolina at Chapel Hill, Chapel Hill, NC, USA**
**www.med.unc.edu/infdis/ideel**
**References:** PMID:29246158, PMID:15132750, PMID:15273102, https://www.ncbi.nlm.nih.gov/pmc/articles/PMC11370457/

**Introduction:** This is a multiplex PCR protocol for simultaneously amplifying *AMA1*, *Heome*, *CRT*, *MDR1*, *DHFR* and *K13 from Chelex-extracted Dried Blood Spot (DBS) samples*.

The PCR primers were adopted from:
- AMA1 - PMID:29246158
- CRT - designed for this protocol
- MDR1 - PMID:15132750
    - MDR184 designed for this protocol
- DHFR & DHPS - PMID:15273102
- K13 – designed for this protocol
- HeOME: https://www.ncbi.nlm.nih.gov/pmc/articles/PMC11370457/

1. **Materials:**
    **1.1 List of primers and expected amplicon lengths based on lab-cultured 3D7 isolate.**

| Gene | Primer Name | Primer Sequence (5'-3') |
|---|---|---|
| *AMA1* | AMA1_F | CCATCAGGGAAATGTCCAGT |
| | AMA1_R | TTTCCTGCATGTCTTGAACA |
| *CRT* | PFCRT_F_v2 | GGTGGAGGTTCTTGTCTTGG |
| | PFCRT_R_v2 | AGTTGTGAGTTTCGGATGTTACA |
| *MDR1* | MDR1_86_F | TGTATGTGCTGTATTATCAGGAGGAAC |
| | MDR1_86_R | AATTGTACTAAACCTATAGATACTAATGATAATATTATAGG |
| | MDR1_184_F_v2 | GTGAGTTCAGGAATTGGTACGA |
| | MDR1_184_R_v2 | GCCTCTTCTATAATGGACATGGT |
| | MDR1_1034_F | AAAAAGAAGAATTATTGTAAATGCAGCTT |
| | MDR1_1034_R | GGATCCAAACCAATAGGCAAAA |
| *DHFR* | DHFR_51_59_F | TGAGGTTTTTAATAACTACACATTTAGAGGTCT |
| | DHFR_51_59_R | TATCATTTACATTATCCACAGTTTCTTTGTT |
| | DHFR_108_F | TGGATAATGTAAATGATATGCCTAATTCTAA |
| | DHFR_108_R | AATCTTCTTTTTTTAAGGTTCTAGACAATATAACA |
| *DHPS* | DHPS_437_F | TGAAATGATAAATGAAGGTGCTAGTGT |
| | DHPS_437_R | AATACAGGTACTACTAAATCTCTTTCACTAATTTTT |
| | DHPS_540_F | AATGCATAAAAGAGGAAATCCACAT |
| | DHPS_540_R | TCGCAAATCCTAATCCAATATCAA |
| | DHPS_581_F | CCTCGTTATAGGATACTATTTGATATTGGAT |
| | DHPS_581_R | TGGGCAATAAATCTTTTTCTTGAATA |
| | DHPS_613_F | TGGATTAGGATTTGCGAAGAAAC |
| | DHPS_613_R | GTTGTGTATTTATTACAACATTTTGATCATTC |
| *K13* | K13-A-432-466_F | GAAAGTGAAGCCTTGTTGAAAGAAG |
| | K13-A-432-466_R | GTACACATACGCCAGCATTGTTG |
| | K13-B-461-531_F2 | TGATGGTGTAGAATATTTAAATTCGATG |
| | K13-B-461-531_R | CTACCATTTGACGTAACACCACA |

| | | |
|---|---|---|
| | K13-C-513-570_F | TTTGAAACTGAGGTGTATGATCG |
| | K13-C-513-570_R | GCTGATGATCTAGGGGTATTCAA |
| | K13-F-567-630_F | TGGCACCTTTGAATACCCCT |
| | K13-F-567-630_R | AGGTAATTAAAAGCTGCTCCTGA |
| | K13-G-660-709_F | TGGCAATTTCTAAATGGTGTACC |
| | K13-G-660-709_R | GCCAAGCTGCCATTCATTTG |
| *Heome* | HeOME-A F | TTTTAGTTTCGGTATTTTGTGTTCCTCTT |
| | HeOME-A R | AAGAAATTTATCAGAGTTACAAAAGGGAAATC |
| | HeOME-B F | TTTATCCTTATCATTATTTCCATCATTTTCTGG |
| | HeOME-B R | AAAAATAAAAGGAACAATGTAATGGTTGAAAA |
| | HeOME-C F | TCCCGAAAACATATCACTAGATCCAT |
| | HeOME-C R | AAAAATTAAACATGATGCCACATTTTAGTAGT |
| | HeOME-D F | AGCTATCATTACATGCTGACACAATAT |
| | HeOME-D R | GATTAGTTGTGGAGATGATAAAACTATCAAATT |
| | HeOME-E F | AATTTTCTTATATAACCTAAGTTGATGACTTGG |
| | HeOME-E R | AGAACAGATGAAGTAACTACTCGATTAAATGA |
| | HeOME-F F | ATCTTTTTCGTTGTATGTGCATAATCA |
| | HeOME-F R | AAACATAATTCTAATGATATTGACCTTGTGCA |
| | HeOME-G F | ACATTCACACAAATAGAAAAATCTTCATTTTTC |
| | HeOME-G R | ATCAATAATCAAAATCATGATAACAACCAATTT |
| | HeOME-H F | AATAACTTAAATAAAAATATGGACGGCTCC |
| | HeOME-H R | GACATTCTTTCAATGCTTCCGAAA |

| | **Chr** | **fullStart** | **fullStop** | **Size** | **He** |
|---|---|---|---|---|---|
| **AMA1** | 11 | 1294287 | 1294523 | 236 | |
| **DHFR-108** | 4 | 748343 | 748493 | 150 | |
| **DHFR-51-59** | 4 | 748173 | 748361 | 188 | |
| **DHPS-436-437** | 8 | 549631 | 549742 | 111 | |
| **DHPS-540** | 8 | 549958 | 550119 | 161 | |
| **DHPS-581** | 8 | 550076 | 550216 | 140 | |
| **DHPS-613** | 8 | 550102 | 550254 | 152 | |
| **Heome-A** | 4 | 115454 | 115684 | 230 | 0.842943 |
| **Heome-B** | 5 | 213026 | 213251 | 225 | 0.837785 |
| **Heome-C** | 14 | 771406 | 771633 | 227 | 0.789254 |
| **Heome-D** | 13 | 2545064 | 2545291 | 227 | 0.77364 |
| **Heome-E** | 8 | 1344617 | 1344845 | 228 | 0.715463 |
| **Heome-F** | 11 | 1816151 | 1816378 | 227 | 0.703947 |
| **Heome-G** | 13 | 815861 | 816091 | 230 | 0.607549 |
| **Heome-H** | 11 | 408542 | 408771 | 229 | 0.605662 |
| **K13-A** | 13 | 1725576 | 1725728 | 152 | |
| **K13-B** | 13 | 1725381 | 1725645 | 264 | |
| **K13-C** | 13 | 1725264 | 1725482 | 218 | |
| **K13-F** | 13 | 1725105 | 1725295 | 190 | |
| **K13-G** | 13 | 1724870 | 1725020 | 150 | |
| **MDR1-1034** | 5 | 960948 | 961039 | 91 | |
| **MDR1-184** | 5 | 958372 | 958599 | 227 | |
| **MDR1-86** | 5 | 958071 | 958206 | 135 | |
| **CRT** | 7 | 403503 | 403693 | 190 | |

#### 1.2 Primer pool ratios

- Mix the primers above ***(both forward and reverse)*** into pools 1 and 2 in the volumes below.

| Primer | Pool 1 Volume | Primer | Pool 2 Volume |
|---|---|---|---|
| AMA1 | 20 | PFCRT_v2 | 12 |
| MDR1-86 | 20 | MDR1-184_v2 | 12 |
| MDR1-1034 | 15 | K13-B | 18 |
| K13-A | 20 | K13-F | 12 |
| K13-C | 25 | DHPS-540 | 24 |
| K13-G | 15 | DHPS-613 | 18 |
| DHPS-436-437 | 15 | DHFR-51-59 | 24 |
| DHPS-581 | 15 | Heome-A | 24 |
| DHFR-108 | 25 | Heome-C | 18 |
| Heome-B | 25 | Heome-E | 18 |
| Heome-D | 20 | Heome-G | 36 |
| Heome-F | 15 | Heome-H | 12 |
| **Total Pool 1 Volume** | **460**μL | **Total Pool 2 Volume** | **456**μL |

#### 1.3 Other reagents

- Nuclease-free water
- QIAGEN Multiplex PCR kit (Cat.206145 or Cat.206143)
- **ZR-96 DNA Clean & Concentrator-5 (Cat.4024**)
- Qubit or PicoGreen supplies for quantifying product (Thermo Fisher, Cat.Q32854 or Cat.Q33232)
- IDEEL iTru library preparation protocol

#### 1.4 Consumables/Supplies

- Nuclease-free PCR microtubes/strips/plates.
- Nuclease-free 1.5uL Eppendorf tubes (for master mixes).
- Cold plates for microtubes and 1.5uL tubes.
- Consumables/Supplies

#### 1.5 Experiment Setup

- Thaw reagents and samples' DNA on ice.
- Clean pipettes and working bench tops with 70% ethanol
- UV-treat the working surface, all equipment, and tubes/strips.
- Briefly vortex and centrifuge each reagent before use.

### 2. Methods:

- Dilute the 100μM stock primers down to 10μM working solutions for each forward and reverse primer.
- Prepare the two primer pools as indicated in **section 1.1** by mixing all forward and reverse primers in the ratios shown in **section 1.2.**
- Prepare the PCR master mix based on the table below. Since there are two primer pools, the recipe below applies to both pools 1 and 2. Additionally, two or more PCR replicates are required per primer pool to resolve PCR and sequencing errors. The resulting setup has four PCR reactions per sample, i.e., two PCR replicates for primer pool 1 and two PCR replicates for primer pool 2.

| Reagents for a 25µl reaction | 1x Reaction |
|---|---|
| Qiagen PCR Master Mix | 12.5 µL |
| Primer Pool Mix⁺ | 2.5 µL |
| Template DNA* | 2.0 µL |
| PCR grade water | 8.0 µL |
| Total Volume: | 25.0 µL |

⁺The “primer pool mix” represents one for pool 1 that is separate from pool 2

*Template DNA volume can be adjusted depending on the concentration of DNA

- For a sample called ***“001”***, a simple naming convention across the two PCR replicates can be:
    - **001-p1-A** & **001-p1-B** - representing the two replicates for primer pool 1.
    - **001-p2-A** & **001-p2-B** - representing the two replicates for primer pool 2.
- Combine the master mix and sample DNA, seal caps/strips/plates, and place in a thermocycler using the following conditions:

| Step | | Temperature | Duration |
|---|---|---|---|
| Initial activation step | | 95°C | 15 min |
| | ***Denaturation*** | ***94°C*** | ***30 sec*** |
| **45 Cycles** | ***Annealing*** | ***60°C*** | ***90 sec*** |
| | ***Extension*** | ***72°C*** | ***90 sec*** |
| Final Extension | | 72°C | 10 min |

- Spot check PCR products from both pools by gel electrophoresis against a 50 bp or 100 bp ladder, using a 2% agarose gel. Expected amplicon sizes are listed above.

3. **PCR purification**

- During the optimisation of this assay, the **ZR-96 DNA Clean & Concentrator-5 (Cat.4024**) was used. Refer to insert protocol for additional guidance.

4. **Product quantification and mixing of P1 and P2**

- If desired, assay the purified products using capillary electrophoresis to ensure that you have the desired PCR fragments (*approx.* 100-300bp) before proceeding to library preparation.
- For small numbers of samples, quantify the DNA using Qubit reagent and mix equal amounts of P1 and P2 product prior to library preparation.
- For larger numbers of samples, quantification by FI on a plate reader is recommended.
    - Make a dilution series of 6 controls containing DNA in TE-Buffer (0.1ng to 100ng)
    - Dilute PicoGreen reagent 1:200 according to manufacturer's protocol in TE buffer.
    - Mix 48µL diluted PicoGreen with 2 µL DNA or control in a black flat-well microplate.
    - Shimmy plate *gently* against palm to level the fluid in the bottom of the well. Cover plate if needed with optical film.
    - Incubate plate for 2 minutes at room temperature in the multimode plate reader before quantifying the fluorescence signal with 485Ex - 535Em filters.
- Use calculated concentration to mix equal ng of P1 and P2 reaction products for each sample replicate to go into library preparation

5. **Library preparation**

- Refer to the IDEEL iTru library preparation protocol.

### Protocol 2. Library Preparation

***iTru Protocol Adapted from Published Materials***

**Infectious Disease Epidemiology and Ecology Lab University of North Carolina at Chapel Hill, Chapel Hill, NC, USA**

**www.med.unc.edu/infdis/ideel**

*Reference: https://peerj.com/articles/7755/#supplemental-information*

#### 1.1 Overview:

This protocol is intended for the preparation of libraries from purified *Pf*-SMARRTT amplicons. The original iTru protocol followed methods for Kapa Hyper Prep Kits using half volumes. This protocol halves the volume again to give ¼ reactions . We're using NEB Modules instead of the actual Kapa Reagents referenced in Adapterama, but the same "Sera-Pure" SpeedBeads. We also omitted the dual size selection. The recipe for SpeedBead Mix is within the supplementary material of this paper.

#### 1.2 Consumables

a. P20 Pipette Tips
b. P200 Pipette Tips
c. Reagent Reservoir (sterile)
d. 96-well PCR plates
e. 50mL Conical Tube (sterile)
f. Foil plate seals
g. Optical plate seals

#### 1.3 Reagents

Library Preparation

a. New England Biotechnology Modules:
   i. End Repair (NEB, Cat.E6050L)
   ii. dA Tailing (NEB, Cat.E6053L)
   iii. T4 Quick DNA Ligase (NEB, Cat.E6056L)
b. 100% Anhydrous Ethanol
c. Molecular Grade Water
d. 1X TE Buffer
e. Elution Buffer (Qiagen, Cat.19086)
f. SpeedBead Mix (see Reference)
g. KAPA HiFi HotStart ReadyMix 2X (KAPA Biosystems, Cat. KK2602)
h. 5uM Stubby iTru Adapter (see Reference)
i. i5 Index Primer Set (**Table S14**)
j. i7 Index Primer Set (**Table S14**)

Library Quantification

Qubit HS dsDNA Kit ( Thermo Fisher, Cat.Q32854), *or*
Quant-iT PicoGreen (Thermo Fisher, Cat.Q33232)

#### 1.4 Other Supplies

DynaMag 96-well Magnetic Plate (Thermo Fisher, 12331D)

Multi-mode Plate Reader (FI Unit)
Thermocycler
P10, P20 and P200 12-tip multi-channel pipettor

**2.0 Protocol**

Notes:

I. Begin from amplified, purified, and combined equimolar *Pf*-SMARRT pools for each sample replicate.
II. Allow SpeedBeads to equalize to room temperature for 30 minutes before using.
III. Keep all reagents and Master Mixes in a cold block or on ice.
IV. Do not vortex NEB enzymes. Gently flick to mix, then spin briefly.
V. Combine Master Mixes immediately before use and return reagents to the freezer promptly.
VI. A list of iTru i5 and i7 Dual Indexing Primers, as well as instructions for synthesizing Stubby Y-yoke Adapters, can be found in the original Adapterama paper within their published Supplemental Materials or an extended list.

---

**2.1 End Repair**

1. Insert 12.5µL of purified amplicons in 5 µL of End Repair Master Mix.

    a. 2.5µL buffer, 1.25µL enzyme, & 1.25µL water (1/4 NEB module recommendations).

2. Cover with foil seal, spin plate briefly, then Incubate in a Thermocycler at 20°C for 30 minutes.

3. Add 50µL of SpeedBeads to the 17.5µL volume, mix by gently pipetting, and incubate at room temperature for 5-15 minutes.
    - Magnet capture (DynaMag 96 well magnet plate).
    - Wash twice with 80µL of 80% EtOH.

---

**2.2 A-Tailing**

4. Aspirate final wash with 200µL pipette tip, then re-aspirate final ethanol drop in bottom of well with 20µL tip.

***If working in 96 well plates, and ethanol is aspirated entirely, the first row or column is likely dry enough for immediate addition of A-tailing Master Mix. If beads appear cracked, they have been allowed to dry for too long.***

5. To dry beads, add 12.5µL A-tailing Master Mix.

    a. 1.25µL buffer, 0.75µL enzyme*, & 10.5µL water.

6. Incubate in a Thermocycler at 30°C for 30 minutes.

7. Add 22.5µL of PEG/NaCl to 12.5µL volume, mix by pipetting, and incubate at room temperature for 5-15 minutes. This step re-binds DNA to beads.
    - Magnet capture & wash twice with 80µL of 80% EtOH.

---

**2.3 Stubby Adapter Ligation**

8. To dry beads add 1.25 µL of 5 µM iTru Y-yoke adapters. Dispense adapters directly onto bead pellets with multichannel. Do not attempt to mix with pipette.

***If using the DynaMag 96-well magnet, it is easiest to add adapters in columns of 8 samples***

9. Add 11.2 µL of Ligation master mix to the adapter-moistened bead. Pipette gently to mix.
    a. 2.5 µL buffer, 1.25 µL enzyme*, & 7.5 µL water.
10. Incubate in a Thermocycler at 20°C for 15 minutes.
11. Add 12.5 µL of PEG/NaCl to the 12.5 µL volume, mix, incubate at room temperature for 5-15 minutes.
    - Magnet capture & EtOH wash twice with 80 µL of 80% EtOH.

*~Here is where you could incorporate size selection if desired~*

12. To dry beads add 12.5 µL of 1X TE Buffer. Incubate at room temperature for 5 minutes. This is the pre-PCR library prep.

***There is no need to capture beads or aspirate supernatants. Adapter ligated sample with beads goes into dual-indexing PCR. Keep pre-PCR Library Prep at 4°C while preparing the Dual-Index PCR Plate.***

---

**2.4 Dual-Index PCR Library Amplification**

13. To a new tube add 12.5 µL of KAPA Hotstart Ready Mix, 1.25 µL of i5 primer, 1.25 µL of i7 primer.
14. Mix 10 µL of the pre-PCR library prep (with speedbeads). Cover with foil seal and spin briefly.
15. Place in thermocycler for 8-12 cycles using the following conditions:

| Step | | Temperature | Duration |
|---|---|---|---|
| Initial activation step | | 98°C | 45 sec |
| | ***Denaturation*** | ***98°C*** | ***15 sec*** |
| **45 Cycles** | ***Annealing*** | ***60°C*** | ***30 sec*** |
| | ***Extension*** | ***72°C*** | ***30 sec*** |
| Final Extension | | 72°C | 1 min |

---

16. Use Qubit or PicoGreen to quantify the PCR product. Capture beads with a magnet and aspirate libraries in supernatant to quantify. The DNA is fully in the TE Buffer at this point. Consider running a HS tape if there is not a significant increase in DNA concentration.

17. If pooling samples for sequencing, combine libraries in equal nanogram amounts by dividing the calculated ng/µL value by the ng of each library desired in the pool.

    Ex. To pool 200 ng of a sample with a concentration of 45ng/µL, divide 200/45 = 4.444~µL

---

18. After pooling libraries, purify with Speedbeads
    a. Add 32 µL of Speedbeads to a 25 µL volume of pooled libraries. Mix gently by pipetting and incubate at room temperature for 5 minutes.
    b. Magnet capture and wash twice with 80 µL of 80% EtOH
    c. To dry beads add 25uL 1X TE or Elution Buffer

19. Libraries are ready to be diluted and loaded onto an Illumina sequencing platform
